# Genetic ancestry shapes context-dependent immune regulation across Indigenous South American populations

**DOI:** 10.64898/2026.10.21.26365630

**Authors:** Cecily Choy, Lucas Vicuña, Maria Patiño, Daniel S. Araujo, Daisy Obispo, Maria Luisa Guevara-Fujita, Susanne Fonseca, Débora Scariot, Cherly Araujo, João Ricardo Guerreiro, Oscar Acosta, Camille Johnson, Kevin Meier, Anne Dumaine, Kelly Williams, Thaynara Lima, Thaissa Rodrigues, André Khayat, Evan Wu, Akansha Gupta, Raúl Aguirre-Gamboa, Bridget Chak, João Barroso-Batista, Fabiola Capetillo, Katherine Marcelain, Ignacia Fuentes, Javiera González, Percibal Beltrán, Yolanda Flores, Teresa Coronel, Josefina Latorre, Joaquín Boutaud, Clayton K. Mickelson, Ryan Langlois, Evan Scott, João Guerreiro, Tábita Hünemeier, Ricardo Fujita, Luis B. Barreiro

## Abstract

Human immune responses remain poorly characterized across Indigenous South American (ISA) populations, limiting our understanding of human immune variation. Here, we generated a single-cell transcriptional atlas of 1.4 million peripheral blood mononuclear cells from 345 individuals representing 13 South American populations, profiled at baseline and following stimulation with lipopolysaccharide, influenza A virus, or *Trypanosoma cruzi*. Genetic ancestry was associated with the expression of a large fraction of the transcriptome, often only after stimulation, and explained more variation than age or sex. On average, *cis*-eQTLs accounted for one-quarter of these ancestry-associated differences. Thousands of regulatory variants were common in ISA populations but rare or absent in European reference datasets. Integrating eQTLs with GWAS revealed stimulation-specific disease-associated regulatory variants and an enrichment of ancestry-associated expression differences among genes with colocalizing signals. Our findings highlight the importance of studying diverse populations across immune contexts to understand the genetic architecture of human immune responses.

## Introduction

Indigenous South American (ISA) populations have a distinctive history that has shaped present-day genetic variation. Following the initial peopling of the Americas at least 15,000 years ago,^1,2^ the first inhabitants dispersed and populated a wide range of environments including the Andean plateau, the Amazon rainforest, and the Pacific coastal plain. Environmental variation and the diverse pathogens that were encountered in each setting imposed selective pressures which shaped immune-related genetic variation.^3,4^ This genetic variation was further reshaped over the last five centuries when European colonization caused the displacement and decline of ISA populations, followed by admixture between Indigenous and European ancestries.^5–7^ Newly introduced pathogens exerted additional selective pressure, reflected in reported signals of post-contact adaptation to smallpox in Andean populations.^8^ Whether shaped primarily by demographic history, genetic drift, natural selection, or some combination of the three, this evolutionary past is expected to have left a functional imprint on immune variation among present-day ISA populations relative to other ancestries. Yet, the genetic variation driving population-specific differences in immune response in ISA populations remains largely uncharacterized.

Characterizing genetic variation in ISA populations may provide a molecular basis for ancestry-associated differences in immune-related diseases, which are documented across South American populations. Hispanic Amerindian ancestry is associated with distinct systemic lupus erythematosus risk loci,^9^ native Peruvian ancestry with tuberculosis progression risk,^10^ and in admixed Colombians, Native American ancestry at the MHC locus is protective against *Trypanosoma cruzi* infection.^11^ These disease associations demonstrate a genetic contribution to immune function in ISA populations, but for immune-related diseases more broadly, most variants identified by genome-wide association studies (GWAS) fall in noncoding regions.^12,13^ This enrichment of noncoding variants suggests gene regulation as a mechanism linking genetic variation to disease. Thus, mapping expression quantitative trait loci (eQTLs), particularly across different stimulation contexts, is essential to identify variants regulating immune genes, many of which appear only after cells are activated.^13–15^

Existing genomics datasets are overwhelmingly biased towards populations of European ancestry.^16–18^ To broaden the representation in genomic datasets, recent studies have generated single-cell immune atlases from peripheral blood mononuclear cells (PBMCs) which include individuals of African^15,19^ and Asian^19–22^ genetic ancestry. However, despite these efforts, South American populations, specifically those of Indigenous ancestry, remain largely underrepresented, limiting our understanding of how ISA-specific genetic variation shapes immune responses.

To address this gap, we present a single-cell transcriptomic atlas of 1.4 million PBMCs, profiled at baseline and after immune stimulation, from 345 individuals representing twelve Indigenous South American populations and one European-ancestry comparison group. These populations, sampled from Brazil, Chile, and Peru, span three distinct ecosystems: the Andes Highlands, the Amazon Basin, and the Pacific Lowlands (the coastal plain west of the Andes). To capture immune responses to distinct classes of pathogen, we stimulated PBMCs with lipopolysaccharide (LPS), a component of Gram-negative bacteria, influenza A virus (IAV), and *T. cruzi*, an intracellular protozoan parasite endemic to the Americas. Using these data, we show that ISA genetic ancestries contribute broadly to variation in the immune response, identify the genetic variants underlying these differences, and link many of them to candidate genes for immune-related diseases, infectious diseases, and hematological traits through colocalization with GWAS risk loci. Together, these results demonstrate the value of incorporating genetically diverse populations into immune genomics.

## Results

### Genetic and single-cell immune profiling of a genetically diverse South American cohort

To characterize genetic and immune variation across South American populations, we established a cohort of 637 individuals from 13 populations living in geographically distinct regions across Brazil, Chile, and Peru (Figure 1A, Supplementary Table 1). This cohort represents twelve Indigenous populations and one Chilean population of mainly European descent. For all individuals, we collected whole blood and isolated PBMCs which were used for genotyping and single-cell RNA-sequencing (scRNA-seq) stimulation experiments. Using the genotype data, we performed principal component analysis (PCA) jointly on our newly genotyped samples and reference panel populations of European and ISA ancestries (Figure S1). Throughout, we use “genetic ancestry” as a conventional shorthand for genetic similarity to reference panel samples, without implying discrete or homogeneous ancestral populations.^23,24^ In the PCA, the first two PCs separated samples into the four broad regional groups represented in our study (“European,” “Andean Highland,” “Pacific Lowland,” and “Amazonian Basin”), capturing the major axes of genetic variation across the continent. PC1, which explained 8.95% of the variance, primarily reflected differences between European and ISA ancestries, with admixed individuals falling along this axis according to their proportion of European ancestry. PC2, which explained 0.86% of the variance, further distinguished the three ISA regions.

**Figure 1.**
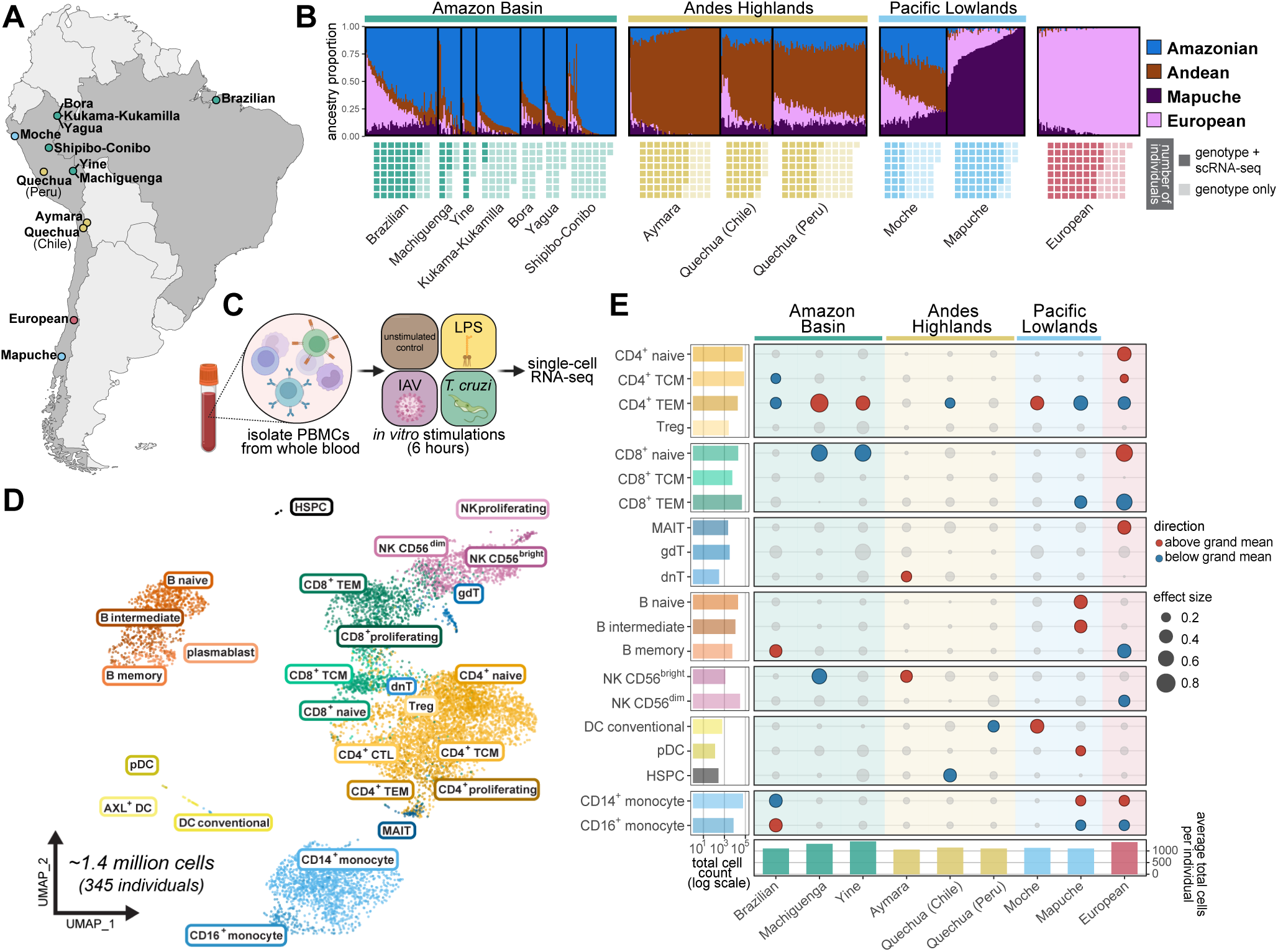
A single-cell RNA-sequencing atlas of baseline and infected PBMCs from 13 populations sampled across South America. (A) Map of populations sampled across Brazil, Chile, and Peru. Indigenous populations are color-coded by the geographical region that they reside in (green = Amazon Basin, yellow = Andes Highlands, blue = Pacific Lowlands). The European Chilean population, though also located in the Pacific Lowlands, is colored pink to distinguish it as a separate non-Indigenous ancestry group. (B) Unsupervised ADMIXTURE genetic ancestry estimates at *K = 4* for all 637 individuals in the cohort. Genetic ancestry components (Amazonian, Andean, Mapuche, European) were named according to the geographic region or population in which each component was maximally represented. Although all individuals in our cohort have genotype information, only a subset of 345 individuals were used for scRNA-seq experiments. (C) Study design for scRNA-seq experiment (created in BioRender, https://BioRender.com/jj3qr4i). (D) Uniform Manifold Approximation and Projection (UMAP) of 1,396,931 baseline, LPS-stimulated, IAV-infected, and *T. cruzi*-infected cells. Abbreviations: CTL, cytotoxic T lymphocyte; DC, dendritic cell; dnT, double-negative T cell; gdT, gamma delta T cell; HSPC, hematopoietic stem and progenitor cell; MAIT, mucosal associated invariant T cell; pDC, plasmacytoid dendritic cell; TCM, central memory T cell; TEM, effector memory T cell. (E) Baseline PBMC compositions differ across populations. Population-level deviations from the grand mean in PBMC proportions were derived from a centered log-ratio-transformed linear model (see Methods). Grey dots indicate non-significant associations (FDR ≥ 0.05; Benjamini-Hochberg correction). Bar charts display total cell counts summed across individuals per cell type (left; log scale) and average total cells per individual for each population (bottom).

ADMIXTURE analysis at *K = 4* further supported the population structure identified by PCA and confirmed that our sampling strategy captured populations with predominantly ISA ancestry. Across the twelve Indigenous populations, individuals derived a mean of 90.0% of their genetic ancestry from ISA components (Figure 1B, Figure S2). This Indigenous ancestry was distributed across three geographically structured components. One was enriched in Andean Highland populations, accounting for 58.5% to 88.4% of the genetic ancestry in Aymara and Quechua individuals, while a second was enriched across populations from the Amazon Basin, where it accounted for 58.6% to 86.9% of inferred genetic ancestry. The European ancestry component, in contrast, was concentrated in the European Chilean population, where it represented 95.2% of genetic ancestry. A fourth component was strongly enriched in the Mapuche, accounting for an average of 81.2% of their inferred genetic ancestry. Consistent with previous studies,^6,25^ this Mapuche-enriched component was genetically distinct from the other South American ancestry components. Moche showed the most heterogeneous genetic ancestry profile among the populations examined. All four components contributed between 16.9% and 33.7% on average, in contrast to the predominance of a single component observed in every other population (Figure 1B).

For a subset of individuals (Figure 1B), we exposed PBMCs to a panel of stimuli representing bacterial, viral, and parasitic stimulations: lipopolysaccharide (LPS; n = 327 paired control-stimulated samples), influenza A virus H1N1 Cal/04/09 (IAV; n = 329 paired samples), and *T. cruzi* (n = 306 paired samples) (Figure 1C). After six hours of stimulation, we performed scRNA-seq on all samples. Following extensive quality-control filtering (see Methods), 1,396,931 high-quality cells from 345 individuals across four conditions were retained for downstream analyses. Cell type annotation, using label transfer from a human PBMC reference dataset,^26^ identified 26 immune cell types at high resolution (Figure 1D). We next asked whether baseline PBMC composition differed across populations by performing compositional data analysis on cell type abundance estimates from unstimulated control samples. In the European population, naive CD4^+^ and CD8^+^ T cells were enriched relative to the overall mean, whereas CD4^+^ and CD8^+^ T effector memory (TEM) cells were depleted (centered log-ratio transformation, FDR < 1 × 10^−4^) (Figure 1E, Supplementary Table 2). This pattern contrasted with two of the three Amazon Basin populations which showed depletion of naive CD8^+^ T cells and enrichment of CD4^+^ TEM cells (FDR < 0.05). Mapuche individuals displayed a different pattern involving several immune lineages. Naive and intermediate B cells, conventional and plasmacytoid dendritic cells, and CD14^+^ monocytes were enriched, whereas CD16^+^ monocytes were depleted (FDR < 0.05). More broadly, with the exception of the CD4^+^ TEM compartment described above, significant differences in cellular composition were generally restricted to a small number of the nine populations examined rather than shared across the cohort. Three cell types, Treg, CD8^+^ T central memory (TCM), and γδ T cells, showed no significant differences in abundance in any population. Together, these results suggest that baseline PBMC composition contributes only modestly to population-level variation in the peripheral immune compartment.

### Transcriptional responses of PBMCs to LPS, IAV, and T. cruzi

We next characterized the transcriptional responses to LPS, IAV, and *T. cruzi* across the five major PBMC cell types, CD4^+^ T cells, CD8^+^ T cells, natural killer (NK) cells, B cells, and monocytes, by collapsing our granular cell type annotations into these broader subsets. PCA of pseudobulk gene expression profiles showed that, depending on cell type, PC1 explained 11.1% to 25.3% of the variance in gene expression (Figure S3). In monocytes, PC1 separated control and *T. cruzi*-infected samples from LPS-stimulated and IAV-infected samples, while PC2 further distinguished these conditions, most clearly separating LPS from IAV (Figure 2A).

**Figure 2.**
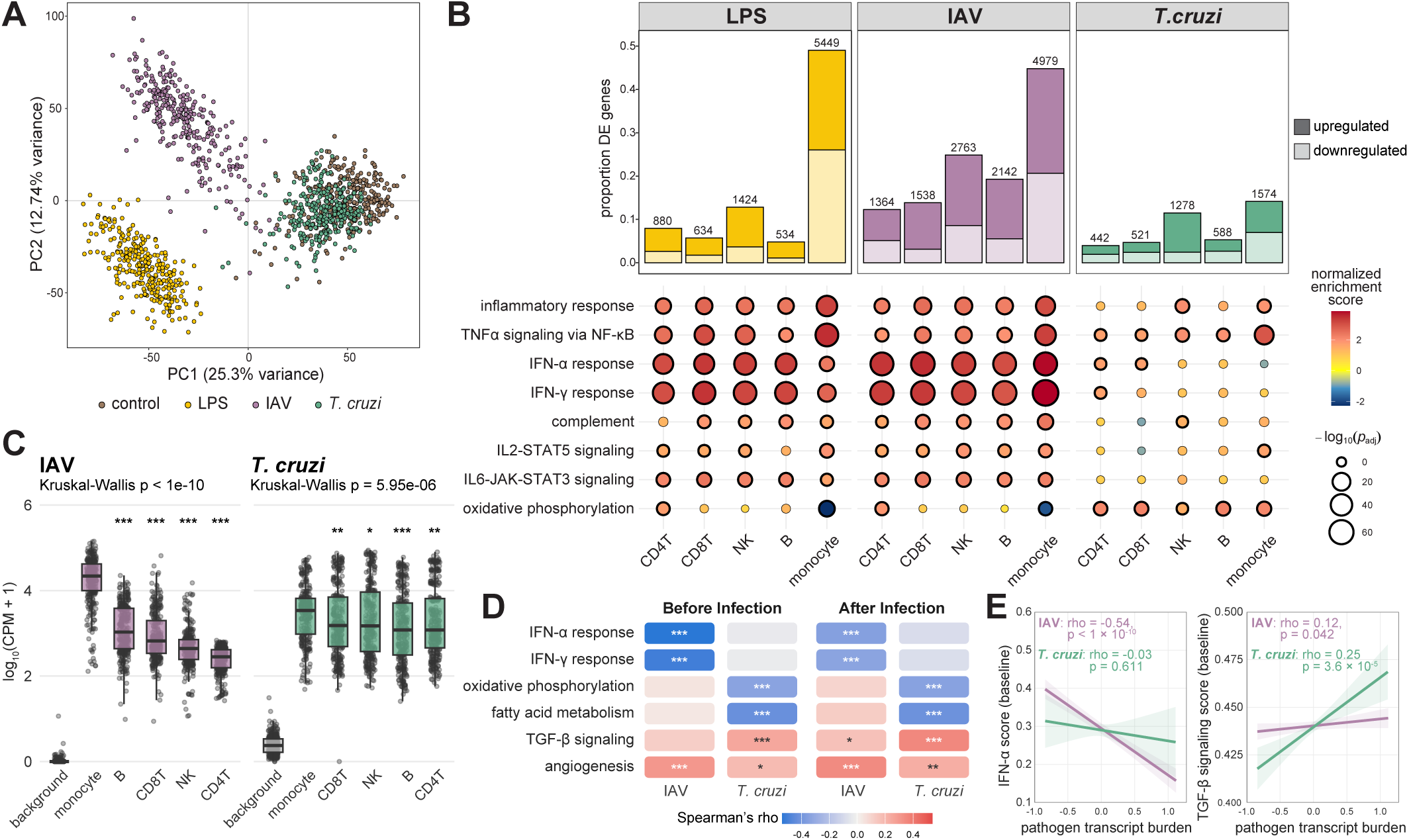
Transcriptional responses to LPS, IAV, and *T. cruzi* stimulation. (A) Principal component analysis of pseudobulk gene expression data from monocytes. (B) Differentially expressed (DE) genes and their enrichment for immune-related pathways. Top: Bar plots showing the number and proportion of DE genes (|log_2_ fold change| > 0.5, FDR < 0.05) for each stimulation condition by cell type. Bottom: Hallmark pathway enrichment analysis for DE genes focusing on immune-related pathways. Only dots outlined in black reach significance (FDR < 0.05). (C) Viral transcript burden is highest in monocytes. Pseudobulk viral transcript abundance (CPM = Counts Per Million, log10-transformed) is shown for each pathogen within its own condition (IAV-condition samples for IAV; *T. cruzi*-condition samples for *T. cruzi*). Cell types are ordered within each condition by descending median. Grey background boxes show the mean pathogen CPM across non-matching conditions (control, LPS, and the other pathogen’s infected condition) pooled across all five cell types. Kruskal-Wallis p-value (top left of each panel) tests for an overall difference across cell types (background excluded); asterisks denote pairwise Wilcoxon rank-sum tests comparing monocytes to each other cell type, Benjamini-Hochberg-adjusted within pathogen (*p<0.05, **p<0.01, ***p<0.001, ns = not significant). (D) Distinct host pathways predict pathogen burden in monocytes, depending on the pathogen. Spearman correlation (rho) in monocytes between select single-sample Hallmark pathway scores and pathogen transcript burden. Spearman’s rho was calculated ‘Before Infection’ (a given individual’s baseline pathway scores from the unstimulated control sample correlated with that same individual’s subsequent titer in the matching infected condition) and ‘After infection’ (pathway score and titer measured concurrently in the same infected-condition sample). Asterisks denote Benjamini-Hochberg-adjusted significance (*p<0.05, **p<0.01, ***p<0.001). (E) Representative examples from the ‘Before Infection’ framing. Baseline IFN-α score and baseline TGF-β signaling score plotted against each individual’s subsequent IAV and *T. cruzi* transcript burden. Lines and shaded bands show the linear fit and 95% confidence interval for each pathogen. Any p-values below 1×10⁻¹⁰ are reported as p < 1×10⁻¹⁰.

We next identified differentially expressed (DE) genes for each condition relative to the matched control (|log2 fold change| > 0.5, FDR < 0.05). LPS and IAV induced broader transcriptional responses than *T. cruzi* across all five cell types, with 534 to 5,449 DE genes following LPS stimulation and 1,364 to 4,979 following IAV infection, compared with 442 to 1,574 following *T. cruzi* infection (Figure 2B, Supplementary Table 3). Monocytes showed the largest response to all three stimuli, with 49.0%, 44.8%, and 14.2% of genes differentially expressed following LPS, IAV, and *T. cruzi* exposure, respectively. In the other four cell types, these proportions ranged from 4.8% to 12.8% for LPS, 12.3% to 24.9% for IAV, and 4.0% to 11.5% for *T. cruzi*. Pathway enrichment analysis further highlighted stimulus-specific responses. These differences were particularly pronounced in monocytes, where LPS induced strong inflammatory and TNFα signaling programs, IAV elicited the strongest interferon responses, and *T. cruzi* was characterized primarily by enrichment of TNFα signaling (Figure 2B, Supplementary Table 4). Together, these results show that both the magnitude and nature of the transcriptional response depend strongly on cell type and stimulus.

Consistent with these transcriptional response patterns, monocytes also harbored the highest pathogen transcript burden for both IAV and *T. cruzi*. Differences across cell types were particularly pronounced for IAV (Kruskal-Wallis p < 1 × 10^−10^), with monocytes showing substantially higher viral transcript levels than the other major PBMC populations. *T. cruzi* transcript burden also differed significantly across cell types (Kruskal-Wallis p = 5.95 × 10^−6^), and although the magnitude of these differences was smaller, monocytes still showed significantly higher parasite transcript levels than the other cell types in pairwise comparisons (Figure 2C).

Given that monocytes showed both the strongest transcriptional response to stimulation and the highest pathogen transcript burden for IAV and *T. cruzi*, we focused the next analysis on this cell type. We asked whether the baseline immune and metabolic state of monocytes was associated with the level of pathogen burden observed after infection. For each donor, we scored the activity of all 50 Hallmark gene sets in the pre-infection control sample and then correlated those scores with pathogen transcript burden measured after infection in the same donor. This allowed us to test whether the cellular state before exposure was associated with the subsequent level of infection. Baseline interferon signaling was negatively associated with IAV burden. In contrast, *T. cruzi* burden showed a different pattern, with negative associations with oxidative phosphorylation and fatty-acid metabolism and positive associations with TGF-β signaling and angiogenesis (Figure 2D). Across the 50 pathways tested, the pathways associated with IAV burden and those associated with *T. cruzi* burden showed little overlap (Supplementary Table 5), indicating that the baseline cellular states linked to pathogen burden differ between the two infections.

Two representative examples illustrate this pathogen specificity (Figure 2E). Higher baseline IFN-α response scores were strongly associated with lower subsequent IAV transcript burden (ρ = −0.54, p < 1 × 10^-10^), with donors carrying a higher resting interferon tone going on to better control the virus, whereas no such association was observed for *T. cruzi* (ρ = −0.03, p = 0.611). In contrast, baseline TGF-β signaling was positively associated with subsequent *T. cruzi* burden (ρ = 0.25, p = 3.6 × 10^-5^), while its association with IAV burden was considerably weaker (ρ = 0.12, p = 0.042) (Figure 2E), consistent with prior work showing that TGF-β signaling promotes *T. cruzi* infection and that TGF-β neutralization reduces parasite burden in cardiac tissue.^27^ Together, these results indicate that distinct, pathogen-specific host programs in monocytes determine subsequent pathogen burden, and that these programs are detectable in the host prior to infection.

### Genetic ancestry-associated differences in the immune response to stimulation

We next sought to identify population differentially expressed (popDE) genes by testing, separately within each cell type and condition, whether gene expression was associated with genetic ancestry at baseline or following LPS, IAV, or *T. cruzi* exposure. Because individuals in our cohort carry varying proportions of four genetic ancestry components, modeling the genetic ancestry proportions directly is complicated by their collinearity. Previous studies of ancestry-associated differences in immune responses have often focused on two-way admixture, most commonly European and African ancestry, and modeled a single genetic ancestry proportion.^14,15^ This approach is less suitable for our cohort, where Amazonian, Andean, Mapuche, and European genetic ancestries each contribute distinct genetic structure. We therefore performed PCA on the *K = 4* genetic ancestry proportions. The resulting three PCs captured 100% of the variance in genetic ancestry proportions (Figure 3A, Figure S4). PC1 explained 48.5% of the variance and primarily separated Andean from European ancestry. PC2 explained 28.9% and primarily separated Mapuche from European ancestry. In contrast, PC3 explained 22.6% and captured variation among Indigenous ancestries, primarily separating Amazonian from Mapuche ancestry. Each individual received a score along each of these three axes. Because the PCs are orthogonal, we used these scores in place of the correlated genetic ancestry proportions to model associations between genetic ancestry and gene expression.

**Figure 3.**
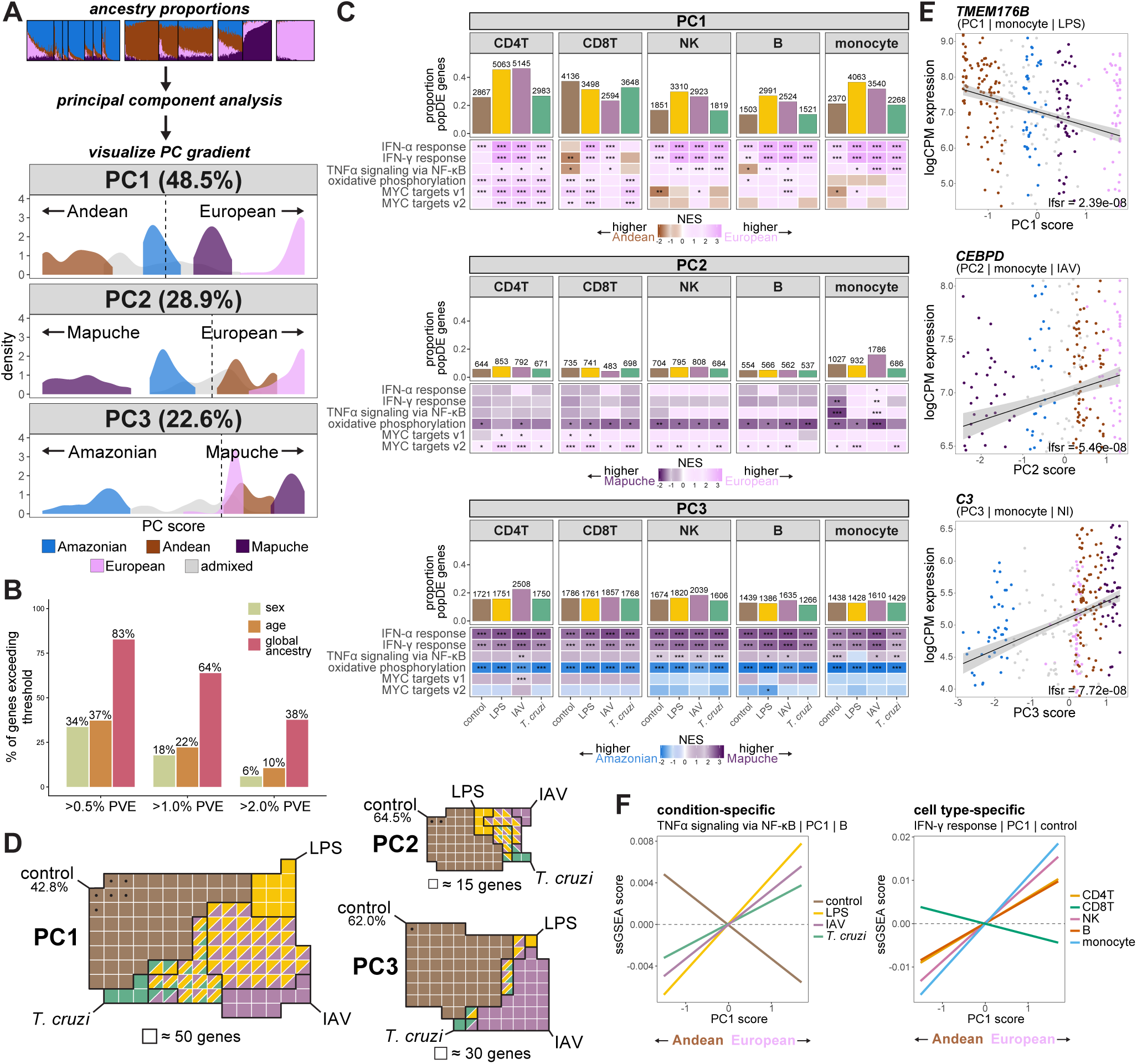
Ancestry-associated differences in gene expression. (A) Calculation of genetic ancestry PC scores from ADMIXTURE genetic ancestry proportions. For principal component analysis and density plot visualization, individuals were categorized by their dominant genetic ancestry component (> 0.60). Individuals whose largest genetic ancestry component did not reach this threshold were categorized as ‘admixed.’ Dashed lines on the density plots indicate where 0 falls on the PC axis. (B) Genetic ancestry explains a substantially larger fraction of gene expression variance than sex or age. Proportion of genes exceeding fixed thresholds of variance explained (PVE) by sex, age, and global genetic ancestry. PVE was estimated per gene, cell type (monocyte, CD4⁺ T, CD8⁺ T, NK, B), and condition (control, LPS, IAV, *T. cruzi*). For each variable, bars show the average percentage of genes with PVE greater than 0.5%, 1.0%, and 2.0% across all five cell types and four conditions. (C) Population differentially expressed (popDE) genes and their enrichment for select Hallmark pathways. Top: Bar plots showing the number and proportion of popDE genes (FDR < 0.05) for each condition by cell type, separately for each PC. Bottom: Heatmap displaying the normalized enrichment scores (NES) from gene set enrichment analysis (GSEA) of popDE genes against Hallmark pathways. Color scale indicates the direction and magnitude of enrichment along each PC’s genetic ancestry axis. Asterisks denote Benjamini-Hochberg-adjusted significance (*p<0.05, **p<0.01, ***p<0.001). (D) Breakdown of popDE genes identified in only stimulation conditions versus at baseline. Venn diagrams for each PC illustrating the number of popDE genes (FDR < 0.05) in baseline (brown), LPS-stimulated (yellow), IAV-infected (purple), and *T. cruzi*-infected (green) conditions. Solid brown cells indicate popDE genes that are identified at baseline and at least one stimulation condition. Brown cells marked with a black dot indicate baseline-specific popDE genes. Grid cell size corresponds to the number of genes as indicated. Percentages indicate the proportion of popDE genes that are found by profiling only the baseline condition. (E) Representative popDE gene examples illustrating ancestry-associated expression along each PC. Scatter plots show logCPM expression of the given gene as a function of PC score. Each point is an individual colored by genetic ancestry group as in Figure 3A. Lines indicate linear fit with 95% confidence interval. (F) Examples of condition-specific and cell type-specific pathway associations with genetic ancestry. All scatter plots show their respective single-sample GSEA scores as a function of the given PC score. Lines indicate linear fit.

Before testing individual genes for associations with genetic ancestry, we asked how much variation in gene expression was associated with genetic ancestry compared with other commonly studied covariates. We used variancePartition,^28^ which estimates the fraction of expression variance explained by each covariate in a linear mixed model. For each gene, we summed the variance explained by the three genetic ancestry PCs to obtain a global estimate of the proportion of expression variance associated with genetic ancestry. At the level of individual genes, the variance explained by genetic ancestry, age, or sex was generally modest, as expected for traits influenced by many genetic, environmental, and technical factors. However, genetic ancestry explained a larger fraction of expression variance across the transcriptome than either age or sex. Across all cell types and conditions, 83% of genes had more than 0.5% of their expression variance explained by genetic ancestry, compared with 34% for sex and 37% for age. At a 2% threshold, the corresponding proportions were 38% for genetic ancestry, 6% for sex, and 10% for age (Figure 3B). This pattern was observed across all five cell types and all four conditions (Figure S5), indicating that genetic ancestry is associated with gene expression across a large fraction of the transcriptome.

We next tested individual genes for associations with genetic ancestry. To improve effect-size estimates and statistical power, we used multivariate adaptive shrinkage (mash),^29^ which borrows information across cell types and conditions based on shared patterns of genetic ancestry effects. Among 11,111 genes tested, we identified 9,197 unique popDE genes at local false sign rate (lfsr) < 0.05 (Supplementary Table 6). PC1 was associated with the largest number of popDE genes (8,735), followed by PC3 (3,363) and PC2 (2,401) (Figure 3C). Many of these associations emerged only after immune stimulation. In CD4⁺ T cells, for example, 57.2% of PC1 popDE genes, 35.5% of PC2 popDE genes, and 38.0% of PC3 popDE genes were detected only after stimulation and would have been missed in control samples alone (Figure 3D). Similar patterns were observed in the other cell types (Supplementary Table 7), showing that immune stimulation reveals ancestry-associated expression differences that are not apparent at baseline.

We then used gene set enrichment analysis (GSEA) to determine whether genes showing ancestry-associated differences in expression were enriched for specific biological pathways (Supplementary Table 8). Along PC1, which primarily separates Andean from European ancestry, genes involved in interferon responses (IFN-α and IFN-γ) and metabolic programs, including oxidative phosphorylation and MYC targets, tended to be more highly expressed toward the European end of the gradient. These patterns were most evident in CD4⁺ T cells, NK cells, and B cells and were stronger after stimulation, in agreement with previous reports of higher interferon pathway activity in individuals with greater European ancestry.^15^ CD8⁺ T cells differed from this pattern, with IFN-γ pathway genes showing the opposite direction at baseline and after *T. cruzi* infection. Along PC3, which separates Amazonian from Mapuche ancestry, interferon and metabolic pathways showed opposing patterns. Interferon-response genes were generally more highly expressed toward the Mapuche end of the gradient, whereas oxidative phosphorylation genes were more highly expressed toward the Amazonian end across most cell types and conditions. Associations along PC2, which separates Mapuche from European ancestry, were generally weaker. Where significant, metabolic pathways such as oxidative phosphorylation tended to show higher expression toward the Mapuche end of the gradient (Figure 3C; representative gene- and pathway-level examples are shown in Figures 3E and 3F).

These data show that genetic ancestry is associated with gene expression across much of the immune transcriptome, but such effects vary largely depending on cell type, and stimulation condition being profiled. Many ancestry-associated expression differences became apparent only after immune stimulation, highlighting the importance of studying genetically diverse populations across multiple immune conditions rather than relying on baseline measurements alone.

### Cis-regulatory variants shape immune gene expression across cell types and stimulation states

To characterize how genetic variants influence gene expression in a cell type- and stimulation-specific manner, we mapped expression quantitative trait loci (eQTLs) separately in all conditions and cell types. For this, we focused on local (*cis*) eQTLs. In total, we identified 6,365 unique genes that had at least one significant eQTL (eGene; lfsr < 0.05) in at least one condition and cell type (Figure 4A, Supplementary Table 9). To assess replication of our results, we compared our baseline eQTLs to eQTLs identified in the OneK1K cohort.^30^ Overall, cell type-matched effect sizes showed high correlation and sign concordance across the five cell types (Pearson’s r = 0.67 - 0.87; concordance = 86.0 - 94.2%) (Figure S6). Furthermore, in every cell type, significant eQTLs were shared between datasets significantly more often than expected by chance (Fisher’s exact test odds ratios = 10.3 - 41.8; p < 1 × 10^⁻10^). Despite this strong overall replication, not all eQTLs replicated across datasets. Some of our non-replicated eQTLs may be explained by differences in minor allele frequency (MAF) between our primarily Indigenous cohort and the primarily European OneK1K cohort. Consistent with this, 28,229 baseline eQTLs detected in our dataset were undetectable in OneK1K specifically because the underlying variants are rare (MAF ≤ 1%) in that cohort, while in our ISA populations these same variants are common, with median allele frequencies ranging from 10.0% to 12.3% across populations (Figure 4B). These thousands of variants represent functional regulatory variation that is invisible to studies focused exclusively on European-ancestry cohorts, underscoring how much population-specific genetic regulation of gene expression remains uncharacterized in the absence of diverse study populations.

**Figure 4.**
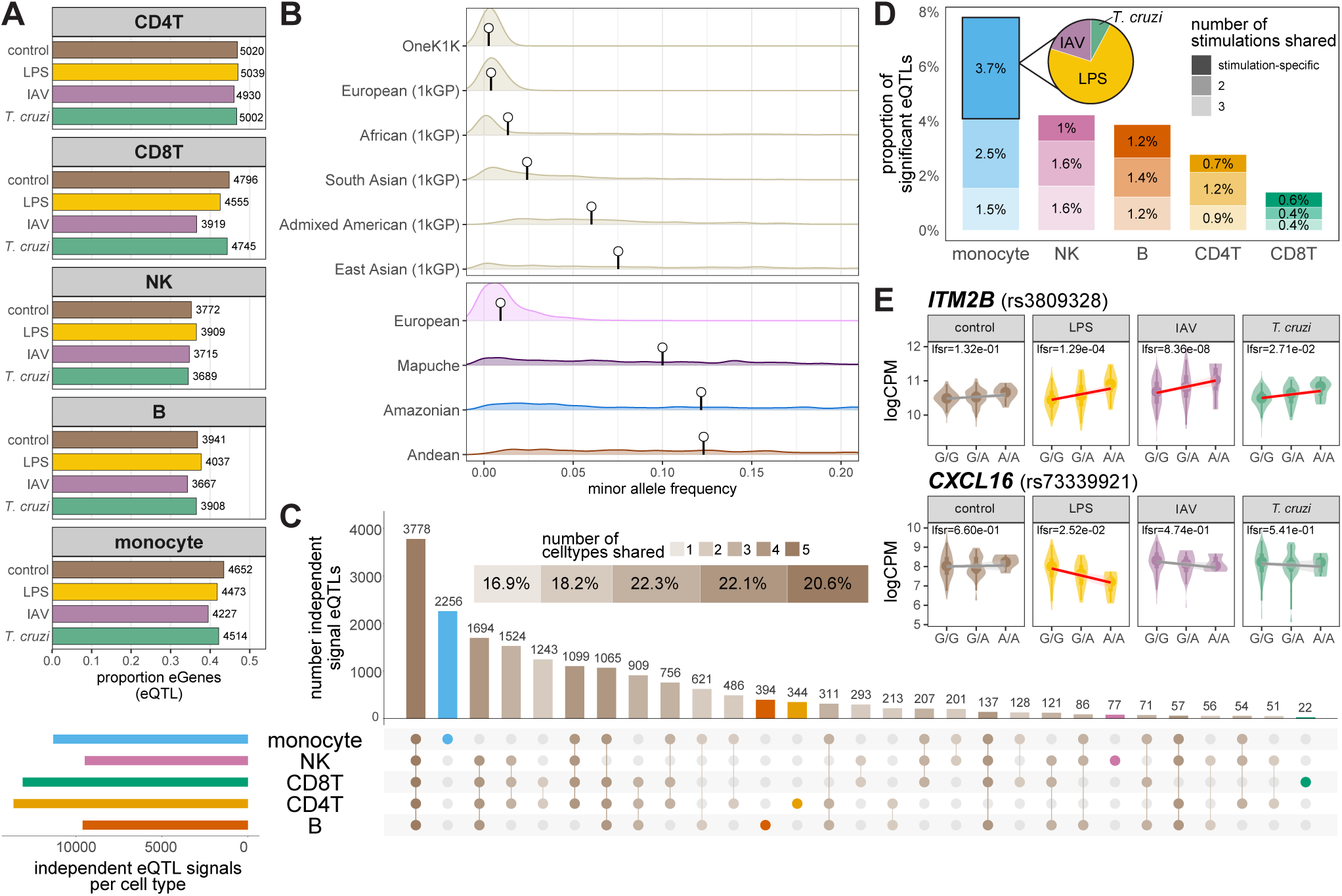
Cis-regulatory variants explain gene expression differences. (A) Proportion and absolute number of eGenes associated with significant eQTLs. Proportion was calculated as the number of eGenes (genes with a significant SNP association, lfsr < 0.05) relative to the total number of genes tested (n = 10,702). (B) Distribution of minor allele frequencies (MAF) for eQTL variants. Variants shown are rare (MAF ≤ 0.01) in the OneK1K reference dataset. Lollipops indicate the median MAF for each distribution. Top: MAF distributions using reference populations from OneK1K and 1000 Genomes Project (1kGP). Bottom: MAF distributions calculated within the genetic ancestry groups of this study. (C) Sharing of baseline eQTLs across cell types. Horizontal bars indicate the total number of independent eQTL signals per cell type; vertical bars indicate the number of independent eQTL signals that are shared among the cell type combination indicated by the connected dots. (D) Sharing of eQTLs across stimulation conditions. Only eQTLs absent at baseline but detected in at least one stimulation condition are included. Inset pie chart shows the breakdown of eQTLs specific to a single stimulation condition in monocytes. (E) Representative examples of eQTLs in monocytes. Plots show logCPM expression by genotype for *IT2MB* (rs3809328), an example of an eQTL shared across all stimulation conditions, and *CXCL16* (rs73339921), an LPS-specific example. Lines indicate linear fit; red lines indicate that the eQTL is significant (lfsr < 0.05) in that condition.

To investigate patterns of eQTL sharing and specificity, we independently examined sharing across cell types and conditions. For eQTLs to be considered shared, we required an lfsr value of 0.05 or less in at least one cell type/condition and 0.1 or less in any additional cell types/conditions. In these additional cell types/conditions, the eQTL effect size was also required to be within a 2-fold magnitude and be concordant in direction relative to the cell type/condition meeting the 0.05 lfsr threshold. In order to assess cell type sharing, we focused on baseline eQTLs and observed that the majority of eQTLs are shared between at least two distinct cell types (83.1%) with 20.6% of eQTLs being shared across all five cell types (Figure 4C). Contrastingly, 16.9% of eQTLs are cell type-specific. Cell type-specific eQTLs were mainly driven by monocytes which had over 2.5 times as many monocyte-specific eQTLs as all other cell types combined. We next assessed eQTL sharing among the stimulation conditions, focusing on eQTLs that were not significant in the baseline condition but were significant in at least one stimulation condition. We found that the percentage of eQTLs found only in stimulation conditions varied across cell types, ranging from 1.4% in CD8^+^ T cells to 7.7% in monocytes (Figure 4D; representative stimulation-specific eQTL examples are shown in Figure 4E).

The existence of eQTLs that are significant only in stimulation conditions suggests that at least some of these variants directly influence the transcriptional response to stimulation, rather than merely crossing a significance threshold due to differences in baseline expression. To test this directly, we mapped response QTLs (reQTLs), defined as QTLs associated with each individual’s change in gene expression upon stimulation relative to their matched baseline control. In total, we identified 850 unique eGenes associated with significant reQTLs (lfsr < 0.05), with monocytes having the largest number of eGenes, consistent with this cell type having the largest response at 6 hours post-stimulation (Figure 5A). Irrespective of cell type, LPS and IAV conditions yielded more reQTLs than *T. cruzi*. To illustrate the range of reQTL condition-sharing patterns we observed, we highlight two examples of reQTLs (Figure 5B). The *SLFN5* locus (rs11658042) displayed a shared reQTL effect across all three stimuli (LPS lfsr = 7.28 × 10^-9^; IAV lfsr = 3.80 × 10^-7^; *T. cruzi* lfsr = 9.64 × 10^-6^). *SLFN5* is an interferon-inducible gene belonging to the Schlafen family of genes, which are involved in immune and interferon responses. The *AAK1* locus (rs58131870) showed a stimulation-specific effect, with a significant association between genotype and change in expression only upon IAV infection (lfsr < 1 × 10^-10^) and not under LPS stimulation (lfsr = 0.93) or *T. cruzi* infection (lfsr = 0.95). *AAK1* encodes AP2-associated kinase 1, a regulator of clathrin-mediated endocytosis,^31^ a pathway used by influenza A virus during cellular entry.^32^ This provides a plausible functional link between genetic variation at this locus and the host response to IAV. We next examined the frequency of the rs58131870 alternate allele across our South American populations (Figures 5C and 5D). Allele frequencies varied across Indigenous populations (MAF = 0.25 - 0.58), but notably, the alternative allele was rare in the European population (MAF = 0.05). This example highlights how ancestrally diverse cohorts can reveal context-specific regulatory variants that are difficult to detect in predominantly European-ancestry populations.

**Figure 5.**
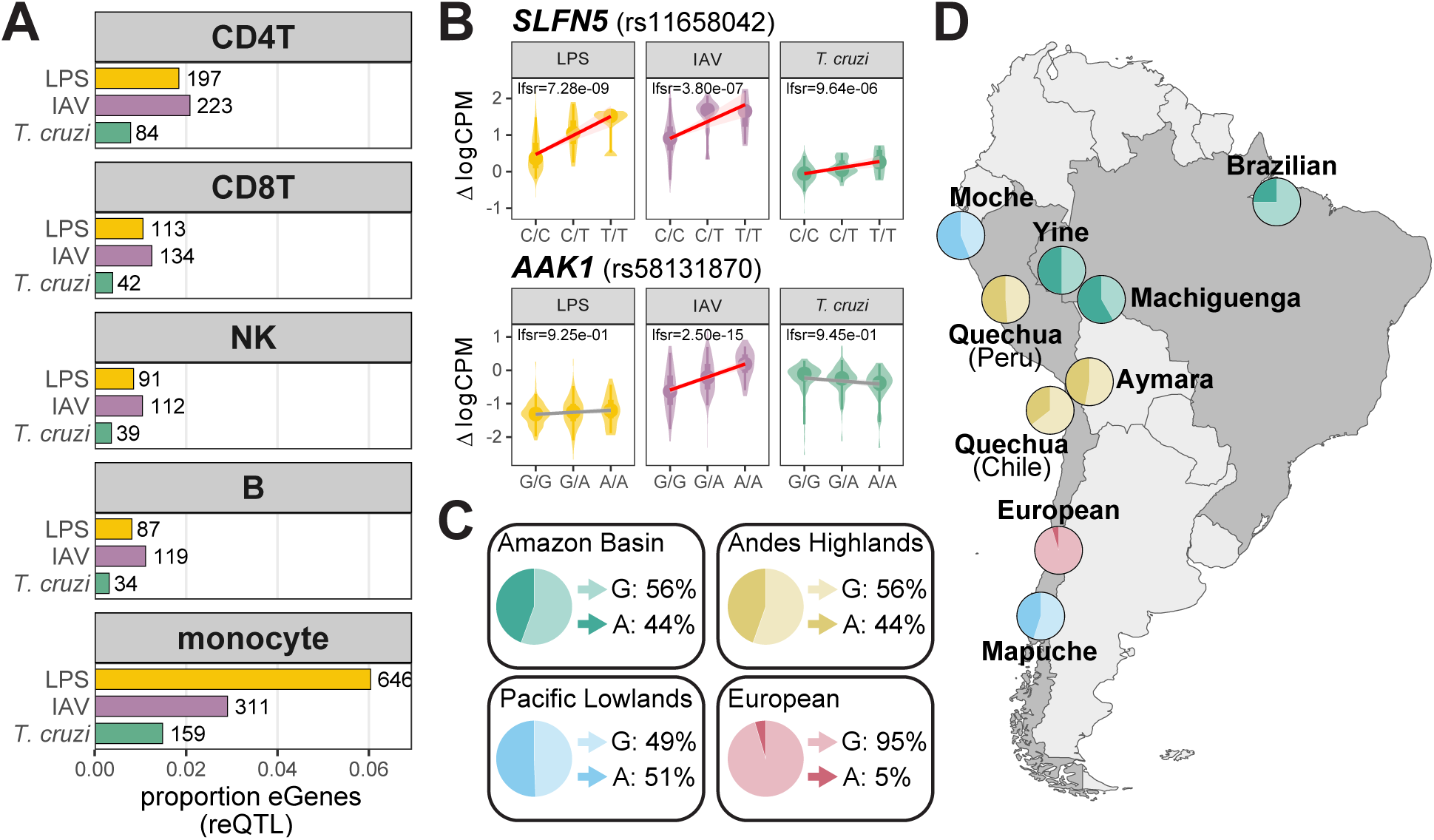
Differences in the transcriptional response to stimulation are explained by *cis*-regulatory variants. (A) Proportion and absolute number of eGenes associated with significant response QTLs (reQTLs). Proportion was calculated as the number of eGenes (genes with a significant SNP association, lfsr < 0.05) relative to the total number of genes tested (n = 10,702). (B) Representative examples of reQTLs in monocytes. Plots show change in expression (Δ logCPM, stimulation condition minus baseline control) by genotype for *SLFN5* (rs11658042), an example of a reQTL shared across all stimulation conditions, and *AAK1* (rs58131870), an IAV-specific example. Lines indicate linear fit; red lines indicate that the reQTL is significant (lfsr < 0.05). (C) Minor allele frequency of *AAK1* rs58131870 variant across sampled geographic groups. (D) Pie charts indicate the frequency of the reference (G) and alternative (A) alleles for each population. Indigenous populations are color-coded by the geographical region that they reside in (green = Amazon Basin, yellow = Andes Highlands, blue = Pacific Lowlands). The European Chilean population, though also located in the Pacific Lowlands, is colored pink to distinguish it as a separate non-Indigenous ancestry group.

### Local genetic variants partially explain ancestry-associated differences in gene expression

After identifying eGenes, we next sought to understand whether these genes under genetic regulation were also likely to be differentially expressed by genetic ancestry. When averaged across cell types, we found that eGenes (lfsr < 0.05) had 1.7- to 3.3-fold higher odds of being classified as popDE genes across PCs and conditions (Figure 6A, Supplementary Table 10). Notably, PC2 consistently displayed higher odds ratios (2.9 - 3.3) compared to PC1 (1.7 - 2.3) and PC3 (1.7 - 1.9), suggesting that PC2 popDE genes are under more genetic control compared to other PCs. Overall, these enrichments point towards ancestry-associated differences in gene expression having a genetic component.

**Figure 6.**
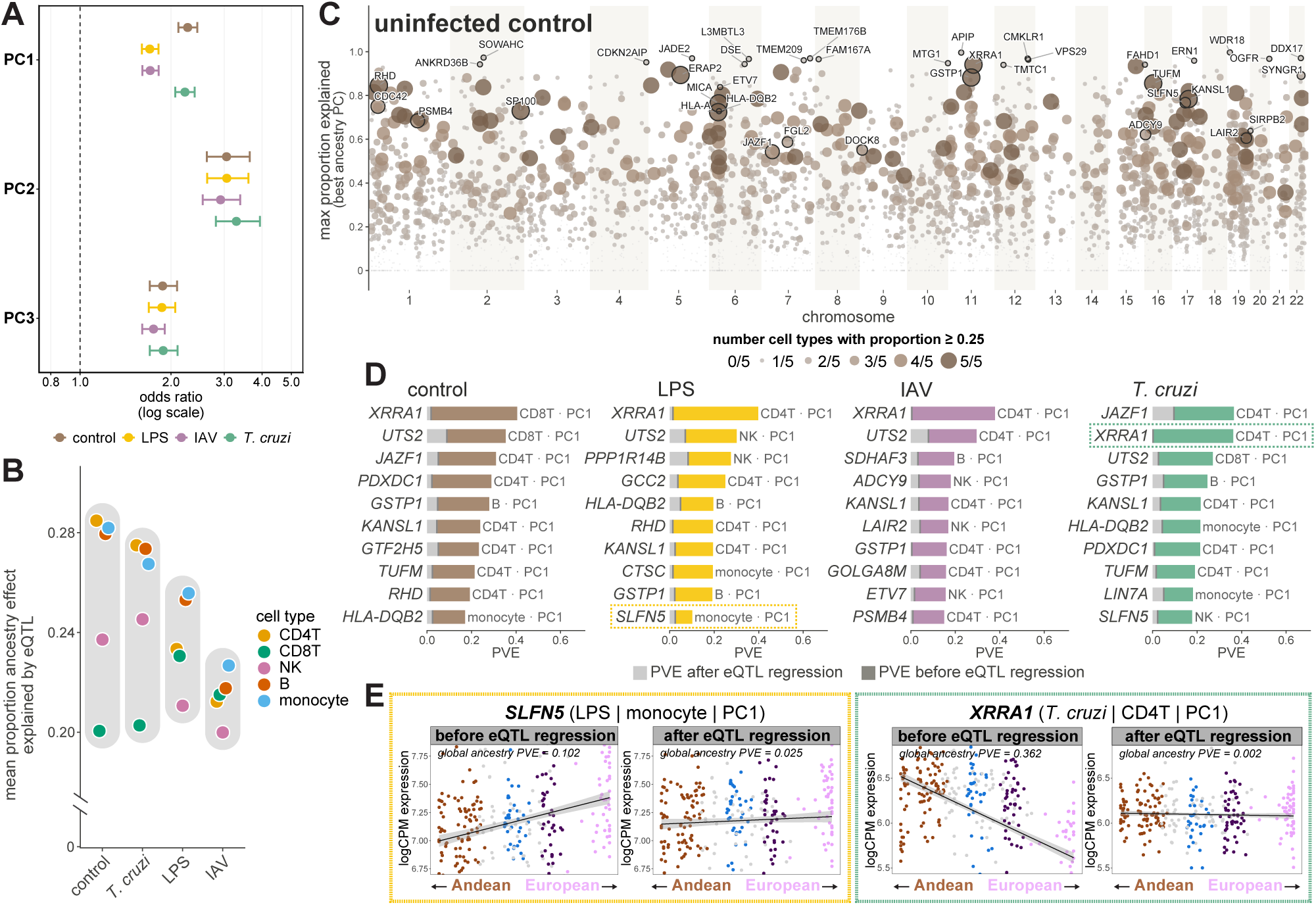
Quantitative genetic contribution to ancestry-associated gene expression. (A) Enrichment of eGenes among popDE genes. For each cell type, condition, and PC, a Fisher’s exact test was used to assess whether popDE genes (lfsr < 0.05) were enriched for eGenes (genes with a significant SNP association, lfsr < 0.05) relative to chance. Odds ratios were pooled across cell types. The dashed vertical line indicates an odds ratio of 1 (no enrichment). (B) Mean proportion of the genetic ancestry effect explained by *cis*-eQTLs, calculated by averaging across all popDE genes with a significant SNP association (lfsr < 0.05). Grey shading highlights the range of values within each condition. The y-axis is broken to omit the region between 0 and 0.20 which contains no data. (C) Manhattan plot showing, for each gene, the maximum proportion of the genetic ancestry effect explained by its eQTL, using the genetic ancestry PC that yields the highest mean proportion explained across cell types. Labeled genes have a proportion explained ≥ 0.5, prioritizing genes with known immune function. (D) Proportion of variance explained (PVE) by genetic ancestry before and after regressing out each gene’s independent eQTL signals. *SLFN5* (monocyte, LPS) and *XRRA1* (CD4T, *T. cruzi*), highlighted with colored boxes, are shown in detail in Figure 5E. (E) Representative examples of ancestry-associated expression before and after accounting for *cis*-eQTL genotype. Each point is an individual colored by genetic ancestry group as in Figure 3A. Lines indicate linear fit. Density plots reproduced from Figure 3A show the distribution of ancestries across the PC1 axis.

To quantify the contribution of genetic variants to ancestry-associated differences in gene expression, we calculated the proportion of genetic ancestry expression variance that is attributed to *cis*-eQTLs for each popDE gene. We did this by extending our variancePartition^28^ framework to additionally include, for each popDE gene, the lead SNP from each of that gene’s independent eQTL signals (identified via fine-mapping and LD pruning, as described in Methods) as covariates, and then recalculated the proportion of expression variance explained by global genetic ancestry. For each popDE gene, we then estimated the proportion of the genetic ancestry effect attributable to *cis*-eQTLs by comparing the ancestry-associated variance explained with and without SNPs included in the model. Among popDE genes with at least one significant independent eQTL (lfsr < 0.05), the mean proportion of the genetic ancestry effect explained by these eQTLs ranged from 0.20 to 0.28 across cell types and conditions (Figure 6B). To confirm that this signal reflected genuine genetic contribution, we compared popDE genes with a significant eQTL to a negative control set consisting of popDE genes which lacked any genetic association. For the control set of genes, we regressed out each gene’s top SNP (by lowest q-value) regardless of significance. The average proportion of the genetic ancestry effect attributable to the regressed variant was substantially lower in this negative control set (mean = 0.080) than in genes with a significant eQTL (mean = 0.237; Figure S7). Together, these results suggest that this signal reflects genuine genetic contribution.

We next asked whether the variants contributing most strongly to ancestry-associated expression differences also showed greater allele frequency differentiation between populations. To address this, we parsed each gene’s eQTL-associated genetic ancestry effect among variants in its fine-mapped credible set and examined their enrichment among genome-wide F_ST_ outliers. Variants accounting for at least 25% of a gene’s genetic ancestry effect were significantly enriched among the top 5% of F_ST_ values, even after stratifying by allele frequency (OR = 1.14 - 2.26; FDR < 0.05 in 17 of 18 population-pair × PC comparisons; Figure S8). These findings further support a role for allele frequency differences at local regulatory variants in shaping ancestry-associated gene expression.

Although *cis*-eQTLs accounted for approximately one-quarter of ancestry-associated expression variance on average, their contribution varied considerably across individual genes. Genes with the largest eQTL contributions were located throughout the genome, including several within the HLA region, and their effect was often shared across multiple cell types (Figure 6C, Supplementary Table 11). To illustrate this relationship at the level of individual genes (Figure 6D), we highlight two representative examples: *SLFN5*, selected for its connection to the stimulation-specific reQTL identified above (Figure 5B), and *XRRA1*, selected because it illustrates how completely a genetic ancestry effect can be attributable to a single local variant (Figure 6E). In monocytes following LPS stimulation, the *cis*-eQTLs associated with *SLFN5* explained a substantial amount (proportion = 0.76) of the gene’s ancestry-associated expression variance. *SLFN5* is an interferon-inducible gene with a response QTL we previously identified, suggesting this variant plays a dual role in both ancestry-associated and stimulation-specific regulation. *XRRA1* represented a more extreme case. In CD4^+^ T cells following *T. cruzi* infection, *cis*-eQTLs for this gene explained 0.995 of the genetic ancestry effect. While such near-complete genetic explanations were rare, they illustrate that ancestry-associated expression can, for some genes, be almost entirely attributable to a single local variant. On average, however, local eQTLs accounted for only about a quarter of the genetic ancestry effect (Figure 6B), indicating that non-genetic or non-local factors also make a substantial contribution to ancestry-associated gene expression.

### Ancestry-associated expression differences are linked to immune disease genes across stimulation contexts

To connect eQTLs to immune-related traits and disease risk, we performed colocalization analysis between our identified eQTLs and GWAS summary statistics for 28 immune-related traits including autoimmune and inflammatory diseases, blood cell traits, and infectious diseases. We considered an eQTL and GWAS signal to colocalize if the posterior probability of a single shared causal variant (PP.H4) was at least 0.8. Across all cell types and conditions, we identified a total of 3,182 colocalizations involving 1,560 unique genes (Figure 7A, Supplementary Table 12). While many colocalizations were shared between baseline and stimulation conditions, a substantial fraction were detected only after stimulation (median 46.9% per trait). For several traits, including rheumatoid arthritis, pulmonary tuberculosis, viral and fungal pneumonia, and sepsis, stimulation conditions yielded more colocalizations than baseline. We next asked whether colocalizations detected in only stimulation conditions were shared across stimuli. Most were specific to a single stimulus (73.1% for autoimmune and inflammatory diseases, 77.0% for hematological traits, and 86.1% for infectious diseases) while only a small minority were shared across all three (5.9%, 5.5%, and 2.8%, respectively) (Figure 7B). Together, these results highlight the importance of immune stimulation for identifying regulatory variants associated with immune-related traits and diseases, many of which would remain undetected in baseline-only analyses.

**Figure 7.**
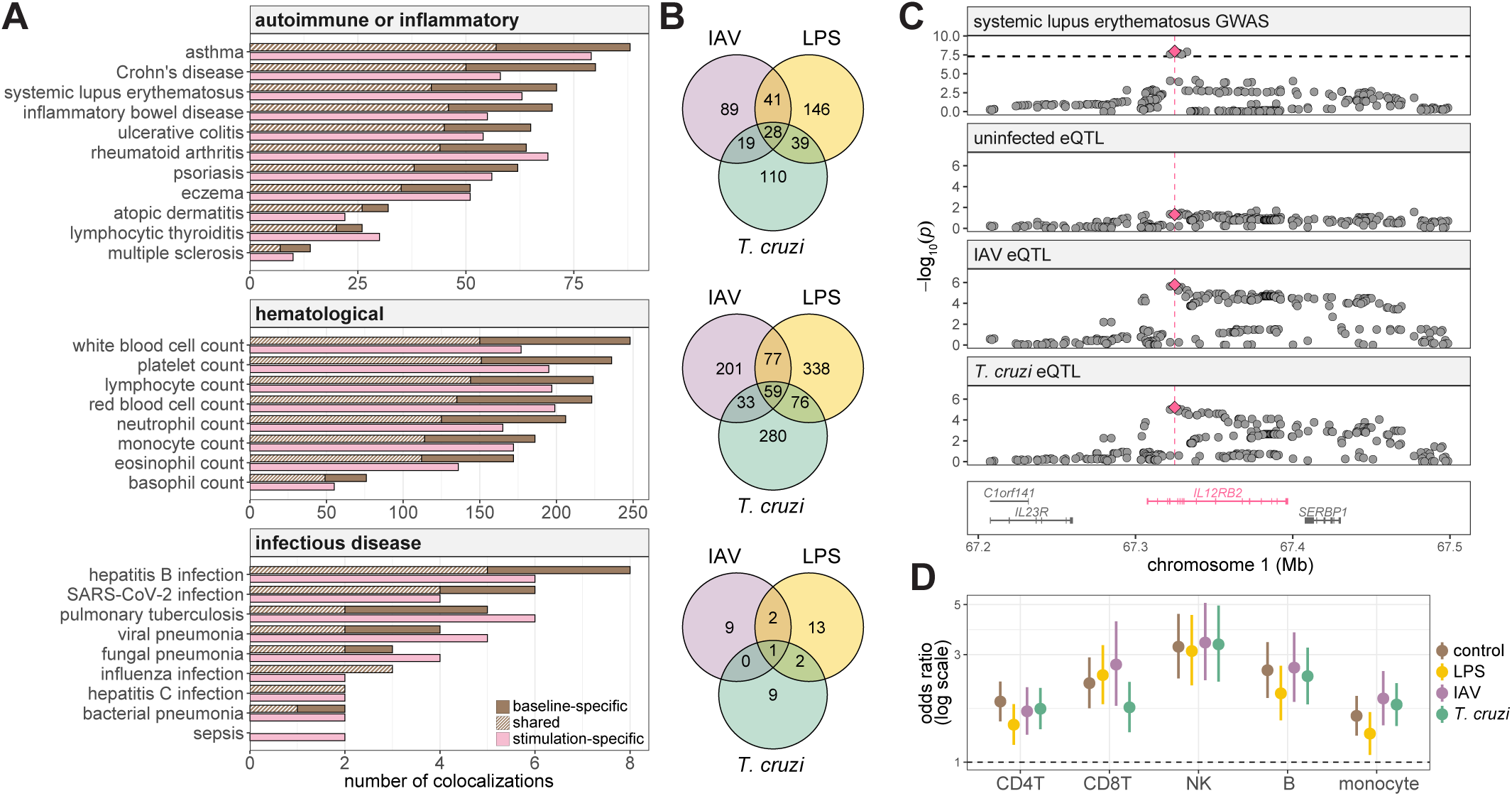
Colocalization of eQTLs with GWAS signals for immune-related traits and diseases. (A) Number of colocalizations between eQTLs and GWAS signals (PP.H4 ≥ 0.8) for 28 immune-related traits. Counts represent the union of colocalizations across all five cell types, with each colocalization counted once if detected in at least one cell type. For each trait, the upper bar shows colocalizations detected in the baseline condition and the lower bar shows those detected in only stimulation conditions. Hatched segments indicate colocalizations shared between baseline and stimulation conditions; solid segments indicate colocalizations specific to baseline (brown) or stimulation (pink) conditions. (B) Overlap of stimulation-specific colocalizations across the three stimulation conditions for autoimmune or inflammatory diseases (top), hematological traits (middle), and infectious diseases (bottom). Counts represent the union across cell type-trait pairs, so a colocalization was assigned to a stimulation condition if it was detected in that condition in at least one cell type. (C) Example of a stimulation-specific colocalization between an *IL12RB2* eQTL in CD4^+^ T cells and a systemic lupus erythematosus (SLE) GWAS signal. The pink diamond and vertical dashed line indicate the lead variant (rs17129789), and the horizontal dashed line in the GWAS panel indicates genome-wide significance (p = 5 × 10^-8^). Gene positions are shown below, with *IL12RB2* highlighted in pink. (D) For each cell type (x-axis) and condition (color), points represent the odds ratio (OR) of a gene being differentially expressed across populations (popDE) among genes with eQTLs colocalizing with at least one of 28 immune-related GWAS traits (coloc.abf PP.H4 > 0.8), compared with eGenes tested for colocalization in the same context but showing no evidence of colocalization. Error bars indicate 95% confidence intervals estimated using Fisher’s exact test. The popDE genes were defined as those showing a significant association with at least one of the first three genetic ancestry PCs (mash lfsr < 0.05). Each gene was counted only once per cell type and condition, regardless of the number of colocalizing GWAS signals. The dashed horizontal line indicates OR = 1. Enrichment was significant across all 20 cell type-condition combinations after Benjamini–Hochberg correction (FDR < 0.01).

One example of a stimulation-dependent colocalization is *IL12RB2*, which encodes the β2 subunit of the IL-12 receptor, and is a popDE gene in CD4+ T cells for both PC1 and PC3. In CD4^+^ T cells, a systemic lupus erythematosus (SLE) GWAS signal at the *IL12RB2* locus colocalized with an *IL12RB2* eQTL in the IAV (PP.H4 = 0.92) and *T. cruzi* (PP.H4 = 0.94) conditions (Figure 7C). In contrast, there was no colocalization between this GWAS signal and the *IL12RB2* eQTL at baseline (PP.H4 = 0.17). This example illustrates how stimulation-induced eQTLs can reveal regulatory mechanisms for disease-associated variants that would have otherwise been undetected in unstimulated cells.

Having established that immune stimulation reveals additional genetic associations with immune-related traits and diseases, we next asked whether these disease-associated regulatory genes were also enriched for ancestry-associated expression differences. To address this, we intersected popDE genes with genes whose eQTLs colocalized with the 28 immune-related GWAS traits. In every cell type and condition, colocalizing genes were more likely to be popDE than other genes tested for colocalization (OR = 1.34 - 3.39; FDR < 0.01 for all 20 contexts; Figure 7D). Importantly, this enrichment was even stronger when considering only genetically regulated popDE genes, defined as those for which local *cis*-eQTLs accounted for at least 25% of the ancestry-associated expression variance (OR = 1.59 - 4.24; FDR < 0.001 in all 20 contexts; Figure S9). Together, these results demonstrate that genes exhibiting ancestry-associated expression differences, particularly those with a substantial contribution from local genetic variation, are enriched for regulatory associations with immune-related traits and diseases. These findings suggest that genetically driven differences in immune gene expression across populations may contribute to population differences in immune phenotypes and disease risk.

## Discussion

Together, our results provide a broad characterization of ancestry-associated variation in immune gene expression across Indigenous South American (ISA) populations. We show that these differences are widespread across the transcriptome, that many only become apparent after immune stimulation, and that local genetic variation explains a substantial fraction of them. We also identify many regulatory variants that are common in our ISA dataset but rare in predominantly European datasets. Colocalization with GWAS further shows that many regulatory variants linked to immune-related traits are detectable only after stimulation and that genes with colocalizing signals are enriched among those whose expression differs by genetic ancestry.

A remarkable aspect of the study was that genetic ancestry was associated with variations in expression across an unexpectedly large proportion of the transcriptome. Across cell types and experimental conditions, 83% of all genes tested showed a significant association with genetic ancestry in at least one cell type and experimental condition. Even though the effects of genetic ancestry were generally weak for most genes, they were consistently larger and affected more genes across the transcriptome than either sex or age, both of which are commonly considered to be major contributors to individual differences in immune variation. This suggests that genetic ancestry, though rarely considered alongside sex and age in immunological studies, may be at least as important of a source of inter-individual variation in immune gene expression.

Ancestry-associated differences in gene expression were involved in a number of biological programs. Across PC1 and PC3, interferon signaling, including both IFN-α and IFN-γ responses, and oxidative phosphorylation were among the pathways most consistently associated with genetic ancestry across cell types and conditions (Figure 3C). These same programs, interferon activity and oxidative/metabolic state, also emerged independently as the strongest predictors of pathogen-specific transcript burden in our infection experiments (Figure 2D and 2E). This suggests that the programs showing the clearest ancestry-related differences are also among those most relevant for shaping how monocytes respond to pathogen exposure. Similar ancestry-associated differences in interferon-related pathways have also been reported in independent comparisons of individuals of European and African ancestry, both in circulating immune cells and in macrophages, where these differences remain detectable even after accounting for *cis*-regulatory genetic variation.^15,33^

However, genetic ancestry is not a biological mechanism. It is a summary of correlated genetic structure among populations. In our study, the populations also varied in geography, lifestyle, diet, altitude, infection history, microbiome exposure, and other factors that were not fully characterized. An association between genetic ancestry and gene expression therefore does not necessarily indicate a direct causal effect of genetic variation. To begin separating these effects, we asked how much of the ancestry-associated signal could be attributed to local genetic variation. On average, *cis*-eQTLs accounted for approximately 20%-28% of the ancestry-associated expression variance among genes with significant local genetic effects. The fact that, for most ancestry-associated genes, *cis*-eQTLs explained only part of the ancestry-associated expression differences in our cohort is not unexpected. Our analysis was specifically designed and adequately powered to identify local regulatory effects; conversely, trans-regulatory effects, which are thought to be common, are generally more difficult to identify and typically require larger sample sizes. Thus, many of the ancestry-associated differences we observed are likely attributable to either unmeasured genetically related effects or environmental factors that co-vary with genetic ancestry.

Despite this relatively modest average contribution, the proportion of ancestry-associated expression variance explained by *cis*-eQTLs varied considerably across individual genes. In some cases, the credible region surrounding the gene accounted for nearly all of the ancestry-associated variation in gene expression in at least one cell type or condition. For example, among popDE genes identified using PC1, the principal component axis that differentiates individuals with Andean versus European ancestry, we identified several key immune-associated genes for which genetic ancestry effects were largely genetically controlled. Local genetic variation accounted for approximately 93% of the ancestry-associated variation in *HLA-A* expression, a component of MHC class I, and 85% for *CIITA*, the master regulator of MHC class II expression. Similar contributions were observed for *NFKB1*, a central regulator of inflammatory signaling, and *SOCS1*, a negative regulator of cytokine and JAK-STAT signaling, with *cis*-eQTLs accounting for as much as 81% and 73% of the ancestry-associated variation, respectively.

Contributions were also significant for *SAMHD1* (an antiviral restriction factor) at up to 67%, and *TLR1* (a bacterial pattern-recognition receptor) at up to 55%. These examples illustrate that for a portion of ancestry-associated immune genes involved in antigen presentation, innate sensing and inflammatory signaling, local genetic variation accounts for a substantial proportion of present-day population differences in gene expression. Additionally, many of the *cis*-regulatory variants contributing to these effects are highly differentiated between populations, with F_ST_ values exceeding the 95th percentile across the entire genome. Whether this differentiation results from genetic drift, natural selection or a combination of both is currently unknown, but the population-resolved eQTL catalog generated here provides a useful framework for addressing this question in future studies.

Beyond characterizing the genetic contribution to ancestry-associated expression differences, our study highlights the importance of including underrepresented populations in studies of immune regulation. We provide the first single-cell atlas of immune responses to stimulation across ISA populations, addressing a gap in the field that has repeatedly been identified as one of its most pressing: the overwhelming concentration of genomic studies on individuals of European ancestry.^16,17^ The consequence of this gap is not merely representational. Thousands of the eQTLs we identified involve variants that are common in ISA populations but too rare to detect in a primarily European reference cohort such as OneK1K (Figure 4B). From the perspective of most existing eQTL catalogs, this regulatory variation is therefore largely invisible. Our results provide direct quantitative evidence for what has so far been largely a conceptual argument for increasing population diversity in studies of immune regulation. This limitation also extends to disease genetics. Because most GWAS have focused on individuals of European ancestry, they have limited power to identify disease associations involving variants that are rare in these populations but common elsewhere. As GWAS expand to include more diverse populations, we expect that some of the regulatory variants identified in our study will also be linked to immune-related traits and diseases. Our eQTL catalog therefore provides a valuable resource for identifying and interpreting these associations, further highlighting the importance of including diverse populations in studies of immune regulation and disease susceptibility.

In addition to population diversity, cellular context emerged as another important determinant of our ability to identify ancestry-associated expression differences and their underlying genetic regulation. At the expression level, many associations with genetic ancestry were not detectable at baseline and only emerged after stimulation with LPS, IAV, or *T. cruzi*. At the genetic level, we observed the same general pattern with many eQTLs only detectable upon immune activation. Thus, stimulation revealed two related but distinct forms of regulatory variation: ancestry-associated differences in expression that were specific to particular immune conditions, and genetic effects whose influence on expression changed following stimulation. These results emphasize that the regulatory architecture of the immune system cannot be fully characterized from resting cells alone. Profiling cells across relevant immune states exposes variation that remains hidden at baseline.

Colocalization analyses with GWAS for 28 immune-related traits and diseases further highlighted the importance of immune stimulation for identifying disease-associated regulatory variants. A substantial fraction of colocalizations were detected only after stimulation, and for several traits, including rheumatoid arthritis, pulmonary tuberculosis, viral and fungal pneumonia, and sepsis, stimulated conditions yielded more colocalizations than baseline. Most of these colocalizations were specific to a single stimulus, highlighting the importance of studying genetic regulation across different immune challenges. This illustrates how studies restricted to resting cells may miss regulatory variants relevant to immune-related diseases. Interestingly, genes with colocalizing GWAS signals were also more likely to exhibit ancestry-associated differences in expression, particularly when local genetic variants explained a substantial proportion of these differences. Together, these findings suggest that genetically driven differences in immune gene expression across populations may contribute to population differences in susceptibility to immune-related diseases.

There are several limitations that should be acknowledged. Samples were collected from multiple geographic sites by multiple teams; consequently technical variation associated with sample collection site remains a concern. We mitigated this source of potential batch effects via standardized sample collection methods, centralized stimulation experiments and single-cell library preparations, and the mixing of individuals from different populations across experimental batches. Furthermore, our analyses employed continuous genetic ancestry axes rather than discrete population labels, thereby minimizing overlap between genetic ancestry and sampling location. Despite these precautions, residual technical effects associated with sample collection site cannot be completely ruled out.

A second important challenge is disentangling the genetic and environmental contributions to the ancestry-associated expression differences identified here. One approach would be to study cohorts with more detailed information on pathogen exposure and health history, allowing specific environmental factors to be tested directly. Unfortunately, such studies are especially rare in Indigenous populations where there is generally limited availability of routine health care services and resources for conducting longitudinal health monitoring studies. A complementary strategy would compare individuals with similar genetic ancestry living in different environments (e.g., rural versus urban settings),^34–36^ thus holding genetically-related background constant while varying environmental exposure.

More broadly, our findings show that population diversity and cellular context each reveal regulatory variation that remains poorly represented in existing datasets. Expanding this framework to additional Indigenous and globally diverse populations, while incorporating richer environmental and clinical information, should provide a more complete understanding of the genetic, environmental, and evolutionary forces shaping human immune variation.

## Materials and Methods

### Study participants

This study was conducted under a central protocol approved by the Institutional Review Board at the University of Chicago (ID: IRB19-0728), with local ethical review and oversight of participant recruitment and sample collection obtained in each country: in Chile by the Comité Ético Científico (CEC-Salud) of Pontificia Universidad Católica de Chile (approval ID: 180814024); in Peru by Comité Institucional de Ética of Universidad de San Martin de Porres (ID: 0000325; FWA 00015320); and in Brazil by the Comissão Nacional de Ética em Pesquisa (approval ID: 7107656 and 8273857). A total of 637 individuals from 13 populations across Chile, Peru, and Brazil were recruited for this study, and signed and written informed consent was obtained from each participant prior to participation. Participants were recruited from Indigenous populations in Brazil, Chile, Peru and a comparison population of mainly European descent in Chile. The breakdown by population is as follows: Aymara (Chile, n = 80), Bora (Peru, n = 20), Indigenous Brazilian representing the six major linguistic groups (Arawak, Jê, Karib, Pano, Tukano and Tupi) geographically widespread in the Amazon region (Brazil, n = 64), European (Chile, n = 88), Kukama-Kukamilla (Peru, n = 38), Machiguenga (Peru, n = 20), Mapuche (Chile, n = 67), Moche (Lambayeque Region, Peru, n = 59), Quechua (Chile, n = 45), Quechua (Peru, n = 81), Shipibo-Conibo (Peru, n = 42), Yagua (Peru, n = 20), Yine (Peru, n = 13). Participants ranged in age from 18-74 years (median age = 38) and included 342 females and 294 males. Recruitment and sample collection were conducted in partnership with local health authorities and community representatives, several of whom are coauthors on this study; community members contributed to participant recruitment, experimental work, interpretation of findings, authorship, and the return of results.

### Blood processing and PBMC isolation

Blood was collected into multiple heparin tubes per donor. A 1 mL aliquot of whole blood was set aside for DNA extraction: erythrocytes were lysed using Puregene RBC Lysis Solution (Qiagen, 158106), and DNA was extracted from the remaining leukocytes using the DNeasy Blood and Tissue Kit (Qiagen, 69506), following the manufacturer’s instructions for “Cultured cells”. PBMCs were isolated from whole blood using SepMate-50 (IVD) (STEMCELL Technologies): blood was diluted 1:1 with PBS + 2% FBS and layered onto Ficoll-Paque PREMIUM 1.073 (Cytiva, 17544652), followed by centrifugation at 1200 × g for 10 minutes at room temperature, with the brake on. After density centrifugation, the enriched layer of PBMCs was collected, washed, resuspended in freezing medium (10% DMSO in FBS) at a concentration of 10 million cells/mL, and cryopreserved. Each cryopreservation vial contained between 5-10 million PBMCs.

### Generation of influenza A virus

The influenza A virus strain used for infections, A/California/04/2009 (Cal/04/09), was rescued by plasmid-based transfection of 293T cells (ATCC, CRL-3216) with Cal/04/09 segments cloned into the pDZ vector, following previously established protocols.^37–39^ Twenty-four hours post-transfection, 7.5 × 10⁵ MDCK cells (ATCC, CCL-34, NBL-2) were added directly to the transfected culture in Opti-MEM (ThermoFisher Scientific, 31985062) supplemented with TPCK-treated trypsin (1 μg/mL); over the following two days, additional Opti-MEM containing TPCK trypsin (2 μg/mL, 500 μL) was added. The culture supernatant was harvested one day later, centrifuged to clear cellular debris, and stored at −80°C. This rescued virus was subsequently amplified on MDCK cells to produce a working stock, while mock-conditioned control media were generated in parallel by culturing uninfected MDCK cells for 48–72 hours and harvesting the supernatant.

Viral stocks were titered by plaque assay on MDCK cells to determine viral titer (PFU/mL) for subsequent infection dosing. MDCK cells were infected in infection media (PBS with 10% Ca/Mg, 1% penicillin/streptomycin, 5% BSA) for 1 hour at 37°C, after which the infection media was replaced with an agar overlay (2X MEM, 1 μg/mL TPCK trypsin, 1% DEAE-dextran, 5% NaCO3, 2% Oxoid agar) and cells were incubated for 40 hours at 37°C before fixation in 4% formaldehyde. Plaques were blocked and immunostained for 1 hour at 25°C in 5% milk, using an in-house polyclonal mouse anti-Cal/04/09 primary antibody (1:5000; raised by infecting mice with 5,000 PFU of Cal/04/09 and harvesting serum 30 days post-infection) and a peroxidase-conjugated sheep anti-mouse secondary antibody (1:5000; GE Healthcare, 45001275), visualized with TrueBlue Peroxidase Substrate (Kirkegaard & Perry Laboratories, 5064728) per the manufacturer’s instructions.

### Generation of Trypanosoma cruzi

Infective *T. cruzi* parasites were isolated from the blood of infected mice. For this procedure, 7-8-week-old male BALB/c mice (Jackson Laboratories) were infected via intraperitoneal inoculation with 100,000 trypomastigotes from the *T. cruzi* Tulahuen strain (ATCC, 30266), which were isolated from cell culture. Parasitemia levels were monitored by counting trypomastigotes under a light microscope using the Pizzi-Brenner method^40^ in fresh blood collected via tail vein bleeding. When the parasitemia peak was reached at approximately 7 days post-infection (dpi), blood was collected from the infected mice via cardiac puncture following standard procedures. The collected blood was aseptically added to a monolayer of epithelial cells (LLC-MK2, ATCC CCL-7) previously seeded in DMEM + 10% inactivated FBS in cell culture flasks and incubated at 37°C in 5% CO_2_ and 20% O_2_. After 24 hours, the supernatant containing mouse blood was discarded and replaced with DMEM + 1% penicillin/streptomycin after gentle washes with 0.01 M PBS to remove most of the remaining erythrocytes. The flasks were then re-incubated under the same conditions. At 5 dpi, a few trypomastigotes and amastigotes were observed in the supernatant. The supernatant was collected daily and replaced with fresh DMEM (without supplementation). Trypomastigotes were purified from amastigotes by centrifugation. Briefly, the collected supernatant was centrifuged at 2451-2740 × *g* for 15 minutes, allowing most cells to deposit in the bottom pellet. The tube was then incubated overnight under the same conditions, allowing the motile flagellated parasites (trypomastigotes) to move towards the supernatant. The supernatant was collected, and trypomastigotes were concentrated by centrifugation at 3202 × *g* for 15 minutes. Trypomastigotes were diluted in 3% buffered formalin and counted under a light microscope using a hemacytometer chamber. A suspension of 3×10^6^ trypomastigotes/mL was prepared in DMEM to be used for subsequent infections.

### *In vitro* PBMC stimulations and sample collection

Stimulation experiments were performed on a subset of 349 participant samples completed across 16 batches. The number of individuals processed in each batch varied from 14 to 26 (mean = 23.4; median = 24). Each batch was balanced for samples from different populations to avoid introducing batch effects confounded with genetic ancestry. For all batches, PBMCs were thawed and cells were seeded onto a 24-well plate and cultured overnight in RPMI 1640 supplemented with 10% FBS, 1% Penicillin-Streptomycin (10,000 U/mL) (Gibco, 15140122), and 1% GlutaMAX™ Supplement (Gibco, 35050061). On the morning of the stimulations, cells were first harvested using a 9 mm J-hook cell lifter to gently lift any adhered myeloid cells, then 200,000 PBMCs were plated onto a 48-well plate at a concentration of 1 million cells/mL. For each individual, PBMCs were exposed to four conditions: unstimulated media (negative control), LPS (10 ng/mL), Cal/04/09 IAV (multiplicity of infection (MOI) of 0.5), and *T. cruzi* (1:5 cell:parasite ratio). Cells were then incubated at 37°C in 5% CO_2_ and 20% O_2_ for 6 hours. After the 6-hour incubation period, cells were harvested using a 2.5 mm narrow blade to ensure all myeloid cells were collected. For each batch of donors, and within a given condition, cells were pooled across all individuals in equal proportions, resulting in four total pools (control, LPS, IAV, and *T. cruzi*)

Given that monocytes are expected to exhibit the strongest transcriptional signal among PBMCs at 6 hours post-stimulation, we enriched for monocytes for batches 5-16 using the EasySep™ Human CD14 Positive Selection Kit II (STEMCELL Technologies, 17858). Briefly, monocytes were isolated from PBMCs through immunomagnetic positive selection and added back to the CD14 negative fraction to achieve a target monocyte composition of 35% per pooled condition. Batches 1-4 did not undergo monocyte enrichment and thus reflect native monocyte proportions.

Cell pools were then prepared for single-cell captures using either the Chromium Next GEM Single Cell 5’ HT GEM Kit v2 (10x Genomics, 1000359) for batches 1-9 or the Chromium GEM-X Single Cell 5’ GEM Kit v3 (10x Genomics, 1000701) for batches 10-16. To prevent confounding with genetic ancestry and 10x Genomics chemistry, samples from the same population were distributed across both single-cell capture chemistries. Cells from the multiplexed pool were loaded to target 60,000 cells per each capture. Although this target exceeds 10x Genomics guidelines due to an increased multiplet rate, overloading increases cell capture throughput and any resulting multiplets can be identified and removed through subsequent genetic demultiplexing. After Gel Beads-in-emulsion (GEMs) generation, reverse transcription, cleanup, and cDNA amplification were performed according to the respective 10x Genomics kit protocol.

### Single-cell RNA-sequencing library preparation and sequencing

Following purification of the amplified cDNA, single-cell gene expression libraries were prepared by fragmenting cDNA into segments and selecting for optimal cDNA fragment size. Sequencing and library-identification indexes were added via End Repair, A-tailing, Adaptor Ligation, and Sample Index PCR. Completed libraries were pooled then sequenced on the Illumina NovaSeq X in a 26 x 10 x 10 x 90 bp configuration or a 28 x 10 x 10 x 90 bp configuration, for captures generated with Chromium Next GEM Single Cell 5’ HT GEM Kit v2 or Chromium GEM-X Single Cell 5’ GEM Kit v3, respectively. For each library, a depth of 50,000 mean reads per cell was targeted.

### Low-pass whole genome DNA sequencing and VCF processing

Low-coverage whole-genome sequencing at an average of 1.34x coverage was performed for all study participants, and the resulting FASTQ files were aligned were aligned to the hg38 human reference genome using BWA-MEM2 (v2.2.1) [10.1109/IPDPS.2019.00041]. Genotypes were imputed by GLIMPSE2^41^ using the joint 1000 Genomes Project and Human Genome Diversity Project (1kGP + HGDP) reference panel.^42^ Quality control was performed using bcftools.^43^ First, only biallelic autosomal SNPs were retained using bcftools view (-v snps -m2 -M2 -r chr1–chr22). Imputation quality was then assessed using the genotype posterior probability (GP) field. For each variant, the proportion of individuals with a maximum GP value below 0.9 was computed; variants exceeding this threshold in more than 10% of individuals were excluded using bcftools view -T. Finally, duplicated chromosomal positions were identified and removed, yielding a final analysis-ready dataset of 637 individuals with 65,384,712 SNPs. rsIDs were then annotated by mapping variants against dbSNP (build GCF_000001405.40, GRCh38) using bcftools annotate.

### Estimation of genome-wide admixture levels

For genetic ancestry inference analyses, we applied a minor allele frequency (MAF) filter of 0.05 and extracted only biallelic SNPs. LD pruning was performed using PLINK v1.9 with a sliding window of 50 SNPs, a step size of 5, and a r² threshold of 0.1. Individual genetic ancestry proportions were estimated with ADMIXTURE version 1.3.0^44^ using the unsupervised clustering algorithm on the 110,065 variants across 637 individuals, testing K = 2 through K = 15 ancestral populations with 10 independent replicates per K value. To address label-switching across replicates, individual genetic ancestry proportion estimates (Q matrices) were aligned using CLUMPP v1.1.2.^45^ CLUMPP was run using the LargeKGreedy algorithm (M = 3) with 1,000 random permutations of runs per K (GREEDY_OPTION 2, REPEATS 1000), and the symmetric similarity coefficient G’ (S = 2) was used to evaluate the similarity between pairs of runs. We found almost perfect concordance in genetic ancestry estimates (mean r = 1) across the 10 independent runs. We repeated the genetic ancestry inference using a less stringent MAF threshold of 0.01. Genetic ancestry estimates were highly concordant with those obtained using MAF > 0.05 (r = 0.993), indicating that the inferred population structure was robust to the choice of MAF threshold. Aligned Q matrices were used for downstream visualization and interpretation of population structure using pong.

### Principal component analysis of genomic data

Study genotypes were merged with reference populations from the 1000 Genomes Project (GBR, IBS (GBR, IBS; shown as “British” and “Spaniards” in Figure 1 for readability),^46^ the Human Genome Diversity Project (French, Bergamo Italian, Basque, Surui, Karitiana, Colombian),^47^ Bolivian Aymara,^48^ and Pehuenche and Huilliche from Southern Chile,^25^ resulting in 1,023 individuals from 24 populations. Variants were filtered for MAF ≥ 0.01 and LD-pruned in PLINK v1.9 (--indep-pairwise 50 5 0.2), leaving 56,911 SNPs. Principal components were computed with PLINK --pca; variance explained per component was calculated as the eigenvalue divided by the number of individuals.

### Generation of human-pathogen merged reference genome

A custom reference genome was constructed by merging three genomic references: the human GRCh38 genome combined with the Influenza A virus strain A/California/04/2009 (H1N1) genome, the *Trypanosoma cruzi* CLBrener Non-Esmeraldo-like genome (TriTrypDB release 68), and their corresponding annotation files. The *T. cruzi* CLBrener genome is a hybrid assembly comprising two haplotypes (Esmeraldo-like and Non-Esmeraldo-like) plus unassigned sequences. To select the most appropriate *T. cruzi* reference, we compared gene detection across four reference strains (CLBrener, Dm28c, Esmeraldo-like, and Non-Esmeraldo-like) in experimental batches 1 and 2. The non-Esmeraldo-like haplotype was selected as the reference because it consistently yielded the highest median counts per detected *T. cruzi* gene across both batches (272 and 130, versus 266 and 126 for Esmeraldo-like, 153 and 74 for Dm28c, and 69 and 32 for CL Brener) together with more detected genes than the Esmeraldo-like haplotype (10,634 and 10,521 versus 10,145 and 10,018). The *T. cruzi* FASTA and GFF files were downloaded from TriTrypDB (https://tritrypdb.org). FASTA sequence headers were standardized by removing all characters following the pipe symbol. The GFF file was converted to GTF format using gffread and sorted by contig and genomic coordinates. The GTF was filtered to retain only protein-coding genes using cellranger mkgtf. The three FASTA files and their corresponding GTF files were concatenated, and a unified reference index was built using cellranger mkref (10x Genomics Cell Ranger).

### Single-cell RNA-sequencing mapping, demultiplexing, and initial cell filtering

FASTQ files from each multiplexed capture were mapped to the merged human-IAV-*T. cruzi* reference genome using the multi function from Cell Ranger v9.0.1 (10x Genomics). Pooled captures were demultiplexed using demuxalot (v0.4.1),^49^ a genetic demultiplexing algorithm that assigns individual identities to cells by matching single-cell reads against genotypes from a common variants file (filtered for SNPs in protein-coding regions with MAF > 0.05) containing all individuals in the pool. Cells were successfully assigned to 346 of the 349 individuals in the pool.

Following genetic demultiplexing, Seurat (v5.1.0) was used to filter out cells that did not meet quality control criteria. A total of 2,819,257 cells were captured prior to filtering. For a cell to be retained, it was required to meet all of the following criteria: (1) assigned as a singlet by demuxalot, (2) between 200 and 6,000 unique genes detected (nFeature_RNA), and (3) less than 10% mitochondrial reads. After filtering, 1,952,736 high-quality cells from 345 individuals remained.

### Cell type assignments, clustering, and UMAP analysis

Cell types were assigned following a two-step process: an initial reference mapping step followed by manual inspection of cell types. In Seurat, FindTransferAnchors was used to identify shared anchors between a PBMC single-cell reference dataset^26^ and our query dataset, and TransferData was then used to transfer cell type labels from the reference to each query cell. Cell type labels were transferred at three different resolutions: celltype.l1, celltype.l2, celltype.l3 (ordered from lowest to highest granularity). To exclude cells with conflicting cell type assignments, cells were filtered out if their celltype.l2 or celltype.l3 assignment was not a valid subset of their celltype.l1 assignment. After applying this filtering criteria, 1,876,228 cells remained.

To increase cell assignment confidence, the initial reference mapping was subsequently followed by a manual inspection of canonical cell type markers. Since manual marker inspection is performed at the cluster level, data integration across batches was first necessary to generate clusters. To begin data integration, individual Seurat objects, each representing a unique batch-condition combination, were first merged by condition to get four condition-specific Seurat objects. Batch-specific layers within each Seurat object were then consolidated using JoinLayers. These four condition-specific Seurat objects were then merged into a single Seurat object with four layers representing each condition. Rather than clustering all PBMCs together, which would obscure finer resolution during manual inspection, cells were subsetted by broad cell types defined by the celltype.l1 label (CD4^+^ T cells, CD8^+^ T cells, NK cells, B cells, monocytes, DCs, other T cells, and other).

Within each subsetted cell type, data were normalized (NormalizeData), variable features were identified (FindVariableFeatures), data were scaled while regressing out batch, percent mitochondrial reads, and 10x Genomics chemistry (ScaleData), and dimensionality reduction was performed (RunPCA). The data were then integrated (IntegrateLayers) using a reciprocal PCA-based method called RPCAIntegration. Following integration, a shared nearest neighbors graph was constructed (FindNeigbors, dims = 1:30), clusters were called (FindClusters, resolution = 0.8), and dimensionality reduction was performed via UMAP (RunUMAP, dims = 1:60). Clusters that co-expressed canonical markers for the subsetted cell type alongside markers of other cell types were identified as likely doublets and excluded from downstream analysis. Canonical marker genes used to define cell types include: CD4^+^ T cells: CD3D^+^, CD3E^+^, CD8A^−^; CD8^+^ T cells: CD3D^+^, CD8A^+^; NK cells: CD3D^−^, NKG7^+^, GNLY^+^, KLRD1^+^; B cells: MS4A1^+^, CD79A^+^; monocytes: CD14^+^, LYZ^+^; dendritic cells: HLA-DRA^+^, CST3^+^; other T cells: CD3D^+^, CD4^-^, CD8A^-^(double-negative); TRGV9^+^, TRDV2^+^ (γδ T cells); TRAV1-2^+^, SLC4A10^+^ (MAIT); HSPCs: CD34^+^. This process was repeated for all broad cell types.

Cells passing manual inspection were then integrated together using the same normalization and clustering parameters to generate a final UMAP. Final cell type annotations (celltype.l1_manual and celltype.l2_manual) were assigned to each cell by combining the reference mapping labels with the manual cluster inspection. After all filtering steps, 1,396,931 high-quality cells from 345 individuals and 4 conditions were retained.

For downstream analyses, high-resolution celltype.l2_manual labels were used to perform cell type composition analysis, while all single-cell RNA-sequencing analysis used celltype.l1_manual annotations. Single-cell RNA-sequencing analysis focused on the five most abundant PBMC cell types (CD4^+^ T cells, CD8^+^ T cells, NK cells, B cells, and monocytes) since their cell numbers were sufficient to detect stimulation and genetic ancestry effects. Dendritic cells, hematopoietic stem and progenitor cells (HSPCs), and other T cells (encompassing double-negative, γδ T cells, and MAIT cells) were excluded from downstream analyses as they fell outside the primary scope of this study and were sparsely represented, with median counts per individual-condition of 3, 1, and 8 cells, respectively.

### Immune cell type composition analysis

To investigate variation in peripheral blood immune cell composition across human populations, we performed compositional data analysis on cell type abundance estimates derived from single-cell RNA-sequencing data. Raw cell type counts were first subjected to quality filtering: donors with fewer than 200 total cells were excluded, and populations represented by fewer than 5 donors after filtering were removed from the analysis. This resulted in a final cohort of 320 donors across 9 populations.

To account for monocyte enrichment performed from batch 5 onward, in which CD14⁺ selection was used to increase monocyte representation in the PBMC pool, cell type proportions were computed within two separate compositional spaces: (i) a non-monocyte compartment, in which proportions were calculated relative to total non-monocyte cells only, and (ii) a within-monocyte compartment, in which CD14⁺ and CD16⁺ monocyte proportions were computed relative to total monocytes. Cell types with more than 80% zero counts across donors were considered too sparse for reliable modelling and were excluded. Centered log-ratio (CLR) transformation was applied within each compartment using a sample-specific pseudocount of 1/N, where N represents the total cell count within the respective compartment, corresponding to the smallest biologically meaningful unit of abundance.

For each cell type, we fitted a linear model of the form:

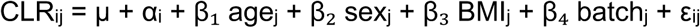

where CLRij is the centered log-ratio abundance of cell type i for donor j, μ is the grand mean, αi is the population effect estimated under sum-to-zero constraints (Σiαi = 0), β₁–β₄ are regression coefficients for the covariates, and εij is the residual error term.

Population was coded using sum-to-zero (effects) contrasts, such that each regression coefficient represents the deviation of a given population from the unweighted grand mean across all populations, without requiring designation of a reference group. Estimated marginal means were recovered for all populations simultaneously using the emmeans package, with effect contrasts applied to extract population-level deviations. Multiple testing correction was applied using the Benjamini-Hochberg procedure across all cell type × population tests simultaneously, and associations with a false discovery rate below 5% were considered statistically significant.

### Calculation of pseudobulk estimates

To create bulk-like expression estimates within samples (defined by a unique individual-condition pair), single-cell counts were aggregated using the AggregateExpression function in Seurat which returns a summed counts matrix across all identified genes. Generation of pseudobulk estimates was performed separately for each of the five main PBMC cell types (CD4^+^ T cells, CD8^+^ T cells, NK cells, B cells, and monocytes). Pseudobulk estimates derived from fewer than 10 cells were excluded, yielding 333, 331, 333, and 312 samples for the unstimulated control, LPS, IAV, and *T. cruzi* conditions, respectively.

### Calculation of capture-corrected expression for downstream modeling

To prepare the raw pseudobulk counts matrices for downstream modeling of stimulation and genetic ancestry effects (see sections below), we first filtered our gene expression matrices to contain only protein coding genes called from the Ensembl database (n = 18,900). We then identified expressed genes for each of the five cell types by only retaining genes whose median count across samples—within that cell type—met our designated cell type-specific thresholds (median log2-counts per million [logCPM] > 1.5 in monocytes, > 2 in CD4^+^ T and B cells, > 2.5 in CD8^+^ T and NK cells). These cell type-specific logCPM cutoffs resulted in the following number of genes per cell type: CD4^+^ T = 9444, CD8^+^ T = 9333, NK = 9542, B = 9779, monocyte = 9450. We then took the union of these cell type-specific gene sets resulting in a combined total of 11,111 genes. Taking the union is a necessary step for downstream mash ^31^ analysis (see “Modeling genetic ancestry effects and integration with mashr” section below) which jointly analyzes effect sizes across cell types and thus requires a common set of genes as input. Using the union ensures that cell type-specific genes that did not meet the expression threshold in other cell types are not excluded from the analysis.

For each cell type-specific gene expression matrix of the union of genes, we calculated normalization factors to scale raw library sizes using calcNormFactors in edgeR (v4.0.16). Through the voom function, raw pseudobulk counts were transformed into logCPM values and the mean-variance relationship was estimated to compute precision weights that were saved for downstream modeling. To account for the technical effect of batch on gene expression, we fit a model (∼ 0 + condition + condition:batch) using the lmFit and eBayes functions. Given our experimental design in which each condition (unstimulated control, LPS-stimulated, IAV-infected, or *T. cruzi*-infected) corresponds to a different lane on the 10x Genomics GEM-X chip, we modeled batch (processing day) as nested within condition; this approach accounts for day-to-day technical variability by lane rather than assuming a shared batch effect across lanes. After fitting the model, the model residuals were obtained using the residuals.MArrayLM function. To preserve the condition effect for downstream modeling of stimulation and genetic ancestry effects, we added back the condition-specific coefficient (i.e., the coefficient corresponding to that sample’s specific condition, estimated per gene) into its residual.

From the batch-corrected gene expression matrix, we constructed two different matrices—each containing a different subset of samples. The first matrix was restricted to individuals with paired control-stimulation samples; any stimulation sample lacking a matched control was dropped. This resulted in 327 paired LPS, 329 paired IAV, and 306 paired *T. cruzi* samples, together representing 333 individuals. This gene expression matrix was used to model stimulation effects (see “Modeling global stimulation effects” section below). The second matrix was restricted to individuals with available BMI metadata, a covariate required for modeling genetic ancestry effects (see “Modeling genetic ancestry effects and integration with mashr” section below). This criterion excluded 3 individuals resulting in 330 control, 328 LPS, 330 IAV, and 310 *T. cruzi* samples.

### Modeling global stimulation effects

To characterize the overall transcriptional response to each stimulation, we modeled the effect of stimulation on gene expression separately for each of the five main cell types (CD4^+^ T cells, CD8^+^ T cells, NK cells, B cells, and monocytes) and each stimulation condition (LPS, IAV, and *T. cruzi*) using the capture-corrected pseudobulk expression estimates described above (“Calculation of capture-corrected expression for downstream modeling”). For each stimulation, samples were subset to the unstimulated condition and the given stimulation condition. To enable a paired design, we retained only donors with samples in both conditions, so that each donor served as their own control. For each cell type and stimulation, we fit the following linear model for each gene using limma,^50^ incorporating the precision weights estimated during voom normalization:

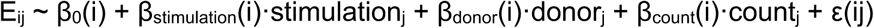

Here, E_ij_ is the capture-corrected expression estimate of gene i for sample j, and stimulation_j_ is an indicator equal to 0 for unstimulated samples and 1 for stimulated samples. Donor was included as a fixed effect, which accounts for all donor-level sources of variation, including genetic ancestry, age, sex, and BMI, without requiring these to be modeled explicitly. Cell count (the number of cells composing each pseudobulk profile) was included as a covariate. Β_stimulation_(i) therefore estimates the within-donor effect of stimulation on gene i’s expression. Models were fit with lmFit followed by empirical Bayes moderation with eBayes.

To control the false discovery rate, we used a permutation-based approach. For each cell type and stimulation, we generated 10 permuted datasets in which condition labels were randomly shuffled within each donor, preserving the paired structure of the data, then refit the model above to each permuted dataset. Empirical p-values were then calculated by comparing the observed −log_10_(p-values) to the null distribution of −log_10_(p-values) pooled across all genes and permutations, using the empPvals function from the qvalue package.^51^ These empirical p-values were converted to q-values using qvalue, and genes were considered significantly differentially expressed in response to stimulation if q < 0.05.

### Detection of intracellular pathogen transcripts

To quantify pathogen transcript burden in each sample, we leveraged reads mapping to the IAV and *T. cruzi* genomes, which were included alongside the human genome in the reference used for alignment (see “Generation of human-pathogen merged reference genome” above). Pathogen transcript burden was calculated separately for each of the five main cell types (CD4^+^ T cells, CD8^+^ T cells, NK cells, B cells, and monocytes) using raw pseudobulk counts prior to gene filtering, so that lowly expressed pathogen genes were retained. For each sample, we summed the counts across all IAV genes and, separately, across all *T. cruzi* genes, and normalized each sum by the sample’s human library size (the total counts across all human genes, excluding pathogen genes) to obtain pathogen counts per million (CPM). CPM values were log10-transformed after adding a pseudocount of 1 (log_10_(CPM + 1)) for visualization.

### Calculation of single-sample pathway scores

To summarize gene expression at the level of biological pathways for each sample, we calculated single-sample pathway scores for each of the 50 Hallmark gene sets from the Molecular Signatures Database (MSigDB)^52^ using single-sample gene set enrichment analysis (ssGSEA), as implemented in the GSVA package.^53^ Scores were calculated using the capture-corrected pseudobulk expression estimates described above (“Calculation of capture-corrected expression for downstream modeling”). ssGSEA was run separately for each combination of the five main PBMC cell types and four conditions (unstimulated, LPS, IAV, and *T. cruzi*), with scores normalized within each run (ssgsea.norm = TRUE). The resulting scores were then combined across conditions within each cell type, yielding a pathway × sample matrix for each cell type.

### Correlation between pathway scores and pathogen transcript burden

To identify cellular pathways associated with pathogen transcript burden, we correlated single-sample pathway scores (see “Calculation of single-sample pathway scores” above) with pathogen transcript burden (see “Detection of intracellular pathogen transcripts” above) separately for IAV and *T. cruzi* in monocytes. For each pathogen, analyses were restricted to samples from the matching infection condition. Because pathway scores were calculated from batch-corrected expression estimates, no further adjustment was applied to them. To account for technical variation in pathogen detection across processing batches, we regressed log_10_(CPM + 1) pathogen transcript burden on batch within each cell type and used the resulting residuals as batch-adjusted measures of pathogen transcript burden.

We examined these relationships in two different framings. First, to assess whether pre-existing pathway activity is associated with subsequent pathogen burden, we correlated pathway scores from each donor’s non-infected, baseline sample with the pathogen transcript burden measured in that donor’s matching infected sample (“before infection” framing). Second, to assess how pathway activity relates to pathogen burden during infection, we correlated pathway scores from each infected sample with the pathogen transcript burden measured in that same sample (“after infection” framing). For each pathway, pathogen, and framing, we calculated the Spearman correlation between pathway scores and batch-adjusted pathogen transcript burden. P-values were corrected for multiple testing across all pathways and cell types within each pathogen and framing using the Benjamini-Hochberg procedure, and correlations with FDR < 0.05 were considered significant.

### Modeling genetic ancestry effects and integration with mashr

To model the effect of genetic ancestry on gene expression and on the transcriptional response to stimulation, we first transformed each individual’s *K = 4* admixture proportions (Amazonian, Andean, Mapuche, and European) into a set of orthogonal genetic ancestry axes. Because admixture proportions are compositional and sum to 1 for each individual, any one of the four could be omitted from a model without loss of information; however, doing so would require an arbitrary choice of which component to treat as the omitted reference and would make the remaining genetic ancestry effects difficult to interpret relative to that omitted group. The remaining proportions would also remain correlated with one another. We therefore performed principal component analysis (PCA) on the *K = 4* admixture proportion matrix (see “Estimation of genome-wide admixture levels” above), which yielded three orthogonal genetic ancestry PCs (PC1–PC3) that together captured 100% of the variance in admixture proportions across individuals. PC scores were mean-centered and scaled to unit variance (z-scored) prior to modeling, as were age. Because the three PCs are orthogonal by construction, all three could be included together as independent predictors of genetic ancestry in the same model, without requiring exclusion of a reference component or introducing collinearity among genetic ancestry terms.

Population differentially expressed (popDE) genes, defined as genes whose expression is associated with genetic ancestry, were identified separately for each of the five main cell types (CD4^+^ T cells, CD8^+^ T cells, NK cells, B cells, and monocytes) using the capture-corrected pseudobulk expression estimates described above (“Calculation of capture-corrected expression for downstream modeling”). For each cell type, we fit the following linear model for each gene using limma,^50^ incorporating the precision weights estimated during voom normalization:

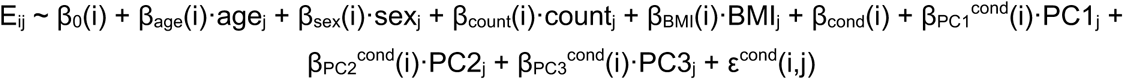

Here, Eij is the capture-corrected expression estimate of gene i for sample j, and condition(j) ∈ {NI, LPS, IAV, *T. cruzi*} denotes the stimulation condition of sample j. Age, sex, cell count (the number of cells composing each pseudobulk profile), and BMI were included as covariates. β_cond_(i) is 0 for the unstimulated reference condition and otherwise estimates the intrinsic, ancestry-independent effect of a given stimulation on gene i’s expression. For each of the three genetic ancestry PCs, a separate coefficient, β_PCkcond_(i), was estimated for every condition, including the unstimulated conditions; this coefficient is the popDE effect of genetic ancestry PC k on gene i’s expression within that specific condition. This model was fit once per cell type, yielding 3 PCs × 4 conditions = 12 popDE effect-size estimates per gene.

To increase power and share information on genetic ancestry effects across cell types and conditions, we applied multivariate adaptive shrinkage (mashr)^29^ separately to each of the three genetic ancestry PCs. For each PC, effect sizes and their standard errors (calculated as the square root of the posterior variance multiplied by the unscaled standard deviation from the limma model fit) were compiled across all five cell types and four conditions into a gene × (cell type– condition) matrix. Correlation structure among conditions was estimated using estimate_null_correlation_simple, and both canonical covariance matrices and data-driven covariance matrices, derived from principal component analysis followed by extreme deconvolution on genes with a significant univariate signal (lfsr < 0.05), were included when fitting the mash model. Posterior effect sizes and local false sign rates (lfsr) were extracted for each genetic ancestry PC and condition. Genes were considered significantly population differentially expressed for a given genetic ancestry PC if the posterior lfsr was below 0.05 in at least one condition.

### Calculation of percent variance explained by model covariates

To quantify the relative contribution of genetic ancestry and other donor characteristics to variation in gene expression, we estimated the percent variance explained (PVE) by each covariate using variancePartition.^28^ Analyses were performed using the capture-corrected pseudobulk expression estimates described above (see “Calculation of capture-corrected expression for downstream modeling”) and were run separately for each combination of the five main PBMC cell types and four conditions. Only samples with complete covariate information, including BMI, were included. For each cell type and condition, we fit the following model for each gene using fitExtractVarPartModel:

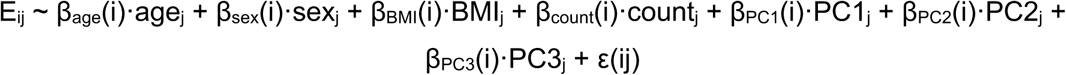

Here, E_ij_ is the capture-corrected expression estimate of gene i for sample j, and age, sex, BMI, cell count, and the three genetic ancestry PCs (see “Modeling genetic ancestry effects and integration with mashr” above) are defined as in the popDE model. Sex was coded numerically (0 = male, 1 = female) so that all covariates were modeled as fixed effects, and the model was therefore fit as a standard linear model. For each gene, this yielded the proportion of expression variance attributable to each covariate, along with the residual variance.

Because cell count reflects a technical property of each pseudobulk profile rather than a donor characteristic, we excluded its contribution and rescaled the remaining variance components, including the residual, so that they summed to 1 for each gene. The resulting values therefore represent the proportion of expression variance not attributable to cell count that is explained by each covariate. Finally, because the three genetic ancestry PCs together capture all variation in admixture proportions, we summed the PVE values for PC1, PC2, and PC3 to obtain a single estimate of the total variance explained by global genetic ancestry for each gene.

### Gene set enrichment analyses

To identify biological pathways associated with transcriptional responses, we performed gene set enrichment analysis (GSEA) using the 50 Hallmark gene sets from MSigDB,^52^ obtained through the msigdbr R package. All analyses were performed using the multilevel algorithm implemented in fgsea (fgseaMultilevel),^54^ considering only gene sets containing between 15 and 500 genes among those tested. To obtain precise p-values for strongly enriched gene sets, the lower bound on p-value estimation was removed (eps = 0). Within each analysis, p-values were corrected for multiple testing across gene sets using the Benjamini-Hochberg procedure, as implemented in fgsea, and gene sets with an adjusted p-value below 0.05 were considered significantly enriched.

To identify pathways enriched among genes responding to stimulation, we performed GSEA separately for each combination of the five main cell types and three stimulation conditions (LPS, IAV, and *T. cruzi*). For each analysis, genes were ranked by the t-statistic for the stimulation effect from the model described above (see “Modeling global stimulation effects”). Gene sets with positive normalized enrichment scores (NES) were therefore enriched among genes upregulated in response to stimulation, and those with negative NES were enriched among genes downregulated in response to stimulation.

To identify pathways enriched among genes associated with genetic ancestry, we performed GSEA separately for each combination of cell type, condition (unstimulated, LPS, IAV, and *T. cruzi*), and genetic ancestry PC (PC1–PC3). For each analysis, genes were ranked by a t-like statistic calculated by dividing the mashr posterior mean by the posterior standard deviation of the popDE effect for the given PC in that condition (see “Modeling genetic ancestry effects and integration with mashr”). This statistic reflects both the direction and the strength of evidence for each genetic ancestry effect, analogous to the t-statistic used for the stimulation response analysis. A positive NES indicates that the gene set is enriched among genes with higher expression in individuals with genetic ancestry toward the positive end of the given PC and lower expression in individuals with genetic ancestry toward the negative end, whereas a negative NES indicates the opposite.

### Quantitative trait loci mapping

To investigate the association between genotype and gene expression, we performed QTL mapping using MatrixEQTL in R.^55^ For each cell type and condition, we adjusted the pseudobulk expression data for batch, age, sex, BMI, and number of cells in the pseudobulk, and quantile-normalized the residuals. In addition to the standard expression QTL mapping, we also performed QTL mapping on the response to stimulation. For this, for each cell type and stimulation condition, we computed the change in expression upon stimulation in comparison to the baseline for every gene. Then, we adjusted the values for batch, age, sex, BMI, number of cells in the baseline pseudobulk, and number of cells in the stimulation pseudobulk, and quantile-normalized the residuals.

To account for cryptic confounders, we performed PCA on the adjusted matrices and identified the best set of PCs by testing up to 20 PCs and choosing the combination that yielded the highest number of eGenes (qvalue < 0.1) in each cell type/condition. The final PC sets are reported below.

| Cell type | Condition | Expression PCs | Response PCs |
| --- | --- | --- | --- |
| B cell | control | 1 to 13 | NA |
| CD4<sup>+</sup> T cell | control | 1 to 16 | NA |
| CD8<sup>+</sup> T cell | control | 1 to 16 | NA |
| Monocytes | control | 1 to 18 | NA |
| NK cells | control | 1 to 17 | NA |
| B cell | IAV | 1 to 20 | 1 to 14 |
| CD4<sup>+</sup> T cell | IAV | 1 to 20 | 1 to 17 |
| CD8<sup>+</sup> T cell | IAV | 1 to 3 | 1 to 13 |
| Monocytes | IAV | 1 to 20 | 1 to 12 |
| NK cells | IAV | 1 to 20 | 1 |
| B cell | LPS | 1 to 11 | 1 to 17 |
| CD4<sup>+</sup> T cell | LPS | 1 to 20 | 1 to 20 |
| CD8<sup>+</sup> T cell | LPS | 1 to 11 | 1 to 5 |
| Monocytes | LPS | 1 to 18 | 1 to 19 |
| NK cells | LPS | 1 to 8 | 1 to 12 |
| B cell | *T. cruzi* | 1 to 14 | 1 |
| CD4<sup>+</sup> T cell | *T. cruzi* | 1 to 19 | 1 to 14 |
| CD8<sup>+</sup> T cell | *T. cruzi* | 1 to 20 | 1 |
| Monocytes | *T. cruzi* | 1 to 19 | 1 to 18 |
| NK cells | *T. cruzi* | 0 | 0 |

To increase our power to identify QTLs, we performed QTL mapping with all individuals in our dataset at once. Thus, to avoid spurious associations due to population structure, we included the first four PCs obtained from a PCA on the genotype data using PC-AiR.^56,57^ For both PC-AiR and QTL mapping, we filtered out genotypes based on minor allele frequency (< 0.01), missingness (> 0.1), or deviation from Hardy-Weinberg equilibrium (p < 1e-6) using plink1.9.^58^ In total, 6,319,656 SNPs passed our filters.

For this analysis, we focused on local (*cis*) QTLs, defined as SNPs within the gene body or 100kb upstream or downstream of the gene transcription start site or transcription end site, respectively. For each cell type and condition, in addition to the standard mapping, we performed 10 additional rounds of mapping with permuted expression matrices. We then used the sets of permuted p-values as our null expectation to estimate FDR using the R package qvalue.^51^ Furthermore, to investigate the existence of multiple, independent signals for a given gene, we fine-mapped our results using susieR.^59–61^

To further improve our QTL mapping results, we used mashr^29^ to leverage sharing of information across cell types and conditions. First, we identified the top SNP for each gene (i.e., the SNP with the lowest qvalue across all conditions and cell types) and defined this set of gene-SNP pairs as our “strong” set. We then randomly sampled 300,000 gene-SNP pairs to fit the mashr model, and applied the fitted model to the strong set to identify genes with a significant QTL effect (local false sign rate < 0.05).

To obtain effect-size estimates for the additional independent signals identified by susieR fine-mapping (see above) within these significant genes, we built a new, expanded strong set and reapplied this same fitted mashr model to it. For each significant gene, we selected the SNP(s) with the highest PIP from each susieR credible set, or, when susieR was unable to identify a credible set, the SNP with the lowest qvalue. Because SNPs within a single credible set can be in strong LD and therefore show similarly high posterior probabilities, we LD-pruned this candidate list (r² < 0.3) to remove this redundancy before designating it as our new “strong” set, as mashr assumes independent signals as input. Applying the same previously fitted model to this new strong set yielded effect-size estimates for each independent signal per gene.

### Expression QTL replication in OneK1K

To assess replication of our expression QTLs (eQTLs), we compared our results with those from OneK1K.^30^ Because the eQTL summary statistics available online were generated with a different pipeline than ours, we instead remapped eQTLs in OneK1K using the same approach described in the “Expression quantitative trait loci mapping” section above, to avoid confounding replication with methodological differences between pipelines. To do so, we obtained imputed genotypes directly from the OneK1K authors and performed build liftover so that coordinates matched the genome build used in our dataset. We also retrieved expression counts from the Human Cell Atlas (see “Data Availability”). Because our own mapping was performed at the level of five broad cell types rather than their finer subsets, we consolidated OneK1K’s more granular cell-type annotations accordingly (e.g., summing “CD14^+^ monocyte” and “CD16^+^ monocyte” counts into a single monocyte category, and its multiple CD4^+^ T cell subtypes into a single CD4^+^ T cell category) so that cell-type granularity matched our dataset.

OneK1K profiled PBMCs at baseline only, without any immune stimulation; we therefore restricted this replication analysis to our unstimulated samples, the only condition directly comparable between the two datasets. After remapping eQTLs in OneK1K, we identified the most significant SNP (lowest qvalue) per gene across all cell types in our unstimulated dataset, and retrieved the corresponding effect sizes and standard errors for the same gene-SNP pairs in both datasets; this became our “strong” set for mashr. We then randomly sampled 300,000 additional gene-SNP pairs, with matched effect sizes and standard errors from both datasets, to fit the mashr model, and applied the fitted model to the strong set.

Finally, using a local false sign rate (lfsr) threshold of 0.05, we performed Fisher’s exact tests to ask whether more eQTLs were significant in both datasets than would be expected by chance. Using the set of eQTLs with an lfsr < 0.05 in our dataset, we further assessed effect size correlation and direction concordance.

### Identification of eQTLs driven by population-enriched genetic variants

To ask whether we identified eQTLs driven by genetic variants unique or enriched in Indigenous South American populations, we first selected eQTLs that were significant in our own control condition (qvalue < 0.05) but rare (minor allele frequency ≤ 0.01) in OneK1K, which represent a cohort predominantly composed of European-ancestry donors. Because these variants are common enough in our populations to provide the statistical power needed to detect an eQTL effect but are rare or largely absent in European-ancestry cohorts such as OneK1K, we would not expect to reliably detect the same effect there. To determine whether the rarity of these candidate variants in OneK1K reflects a broader pattern of ancestry-specific variation rather than a peculiarity of that particular cohort, we retrieved genotype data from the 1000 Genomes Project (1kGP)^62^ and assessed the frequency of these variants across the five 1kGP superpopulations: African, Admixed American, East Asian, European, and South Asian.

### Determination of eQTL sharing across cell types and conditions

To characterize patterns of eQTL sharing, we used the posterior effect sizes and local false sign rates (lfsr) estimated by mashr for each independent eQTL signal (see “Quantitative trait loci mapping” above). Sharing was assessed separately across cell types and across conditions using the same criteria. We first considered an eQTL significant in a given cell type or condition if its lfsr was below 0.05, and restricted analyses to eQTLs that were significant in at least one cell type or condition. Because applying a single significance threshold independently to each context tends to underestimate sharing, we additionally considered an eQTL to be shared with a context in which it did not reach significance if its effect in that context closely matched its effect in the significant contexts. Specifically, for each eQTL we calculated the mean posterior effect size across all contexts in which it was significant and considered the eQTL shared with an additional context if its posterior effect size in that context had the same sign as this mean, was within 2-fold of it in magnitude, and had an lfsr below 0.1. Thus, an eQTL was counted as having an effect in a given cell type or condition if it was either significant there (lfsr < 0.05) or met all three of these criteria.

### Enrichment of eGenes in popDE genes

To assess whether genes under local genetic regulation were more likely to show ancestry-associated expression differences, we tested for enrichment of eGenes among popDE genes. Analyses were restricted to genes tested in both the popDE and eQTL analyses, which excluded mitochondrial and sex-linked genes that were not included in eQTL mapping. For each gene, cell type, and condition, a gene was classified as popDE for a given genetic ancestry PC if its mashr lfsr was less than 0.05 (see “Modeling genetic ancestry effects and integration with mashr” above), and as an eGene if at least one of its independent eQTL signals had a mashr lfsr below 0.05 (see “Quantitative trait loci mapping” above).

For each combination of genetic ancestry PC, condition, and cell type, we constructed a 2 × 2 contingency table of popDE status by eGene status and calculated the log odds ratio. To summarize enrichment across cell types, we averaged the log odds ratios across the five main cell types for each PC and condition and exponentiated the result to obtain an average odds ratio. To estimate uncertainty, we performed a gene-level bootstrap, in which genes were resampled with replacement 2,000 times and the average odds ratio was recalculated for each resample. Resampling was performed at the level of genes, rather than individual gene-cell type-condition observations, and 95% confidence intervals were defined as the 2.5th and 97.5th percentiles of the bootstrap distribution.

To test whether enrichment differed between genetic ancestry PCs, we compared the average log odds ratio for PC2 with those for PC1 and PC3 within each condition. For each comparison, we calculated the difference in average log odds ratios within each bootstrap resample and computed a two-sided bootstrap p-value as twice the proportion of resamples in which the difference fell on the less frequent side of zero, with a pseudocount of 1 added to both the numerator and denominator. P-values were corrected for multiple testing across conditions using the Benjamini-Hochberg procedure.

### Calculation of SNP contribution to percent variance explained by global genetic ancestry

To quantify the extent to which local genetic variants account for ancestry-associated differences in gene expression, we extended the variancePartition framework described above (see “Calculation of percent variance explained by model covariates”) to include eQTL genotypes as additional covariates. Analyses were performed separately for each combination of the five main cell types and four conditions, for every popDE gene in that cell type and condition. For popDE genes with at least one significant eQTL (lfsr < 0.05), we included the lead SNP from each of the gene’s independent eQTL signals (see “Quantitative trait loci mapping”). For popDE genes without a significant eQTL, we included only the gene’s top SNP (lowest q-value), regardless of significance; these genes served as a negative control. Genotypes were coded as 0, 1, or 2 copies of the alternative allele and included as fixed effects, alongside all covariates from the original model (age, sex, BMI, cell count, and genetic ancestry PC1–PC3). As in the original model, the contribution of cell count was removed and the remaining variance components were rescaled to sum to 1, and the variance explained by PC1, PC2, and PC3 was summed to obtain the total variance explained by global genetic ancestry. For each gene, we then estimated the proportion of the genetic ancestry effect attributable to *cis*-eQTLs as (PVE_ancestry,before_ − PVE_ancestry,after_) / PVE_ancestry,before_, where PVE_ancestry,before_ and PVE_ancestry,after_ are the global genetic ancestry PVE from models fit without (before) and with (after) the gene’s SNPs.

For popDE genes with a significant eQTL, we further determined how this genetic ancestry effect was distributed among the gene’s individual independent eQTL signals using Shapley values, which partition the total reduction in genetic ancestry PVE among a gene’s SNPs while accounting for correlation between them. For each gene, we fit a full model including all of the gene’s lead SNPs and a baseline model including none of them. To place all models for a given gene on the same scale, the variance components from every model were divided by a single denominator, defined as the sum of all non-cell count variance components in the full model. The Shapley value for SNP i was then calculated as

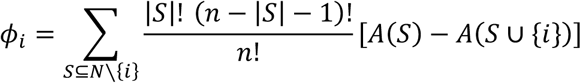

where N is the set of n lead SNPs for the gene, S is a subset of the remaining SNPs, and A(S) is the genetic ancestry PVE from a model including the SNPs in S. Each φ_i_ therefore represents the average reduction in genetic ancestry PVE when SNP i is added to the model, taken across all possible combinations of the gene’s other SNPs, and the Shapley values for a gene sum to the total reduction in genetic ancestry PVE between the baseline and full models. For genes with 10 or fewer SNPs, Shapley values were calculated exactly by fitting all 2^n^ possible subset models. For genes with more than 10 SNPs, Shapley values were approximated by Monte Carlo sampling of 400 random orderings of the gene’s SNPs, adding SNPs one at a time in each ordering and averaging each SNP’s marginal reduction in genetic ancestry PVE across orderings. Because each Shapley value represents the average reduction in genetic ancestry PVE attributable to SNP *i*, the proportion of the genetic ancestry effect attributable to each SNP was calculated as φ_ᵢ_ / PVE_ancestry,before_, analogous to the gene-level calculation.

### Genotype quality control for F_ST_ calculation

Before calculating F_ST_ values per SNP, we removed related individuals up to the second degree. Relatedness among the 639 individuals was assessed using KING^57^ with a kinship degree threshold of 2. Related pairs were classified according to estimated kinship coefficients: duplicate/MZ twins (> 0.354), first-degree relatives (0.177 - 0.354), and second-degree relatives (0.0884 - 0.177). To determine which individuals to remove from related pairs while removing relatedness up to the second degree, we applied the NAToRA algorithm^63^ to select the minimum number of individuals most connected to others. This yielded a dataset of 549 unrelated individuals.

We then performed population-aware MAF filtering on the 549-individual dataset to maximize retention of population-specific variants while removing globally rare variants. Individuals were assigned to four population groups: European (n=88), Mapuche (n=65), Andean (n=232, comprising Aymara, Quechua, and Moche individuals), and Amazonian (n=164, comprising Brazilian, Shipibo-Conibo, Kukama-Kukamilla, Bora, Yagua, Machiguenga, and Yine individuals). For each group, variants with MAF ≥ 0.05 were identified using bcftools filter. The union of the four group-specific site lists was computed, retaining any variant passing the MAF threshold in at least one group (5,934,954 SNPs).

### Calculation of per-SNP F_ST_ scores

To quantify genetic differentiation between ancestry groups, we calculated F_ST_ for all SNPs retained in the VCF detailed above in the “Genotype quality control for selection scans” section. Because these statistics may be biased when including admixed individuals, we performed the analyses separately for the 549 unrelated individuals as well as a subset of 433 individuals with less than 10% European ancestry (and the European Chilean samples), which was re-filtered for SNP MAF > 0.05. In addition, we grouped samples differently across two separate runs using the population (e.g. Aymara, Bora) or population group (e.g. Andean, Amazonian) variables detailed above.

Per-SNP F_ST_ scores were calculated using the Hudson estimator^64^ using Plink.^65^ We supplied the VCFs and sample groupings to the plink2 --fst command with the options method=hudson and report-variants to produce both per-variant and global F_ST_ scores across all pairs of sample groupings. Negative F_ST_ scores indicate more within-population variation than between populations and were therefore set to zero before further analysis.

### Enrichment of F_ST_ outliers among variants explaining ancestry-associated expression

In order to assess if the variants responsible for ancestry-associated expression differences are more differentiated between genetic ancestry groups, we evaluated the per-variant proportion of the genetic ancestry effect as described above, taking the maximum value over genes, cell types, and conditions for each genetic ancestry PC. Variants that explain at least 25% of the genetic ancestry effect and credible-set variants were compared separately against the rest of the variants in the genome-wide FST dataset (5,934,954 SNPs). For each of the six pairs of population groups, F_ST_ outliers were defined as variants with F_ST_ values in the upper 5% of the genome-wide per-SNP F_ST_ distribution for that particular pair. As eQTL variants have a tendency to be composed of common alleles and Hudson F_ST_ relies upon allele frequency, we used a Cochran-Mantel-Haenszel test to examine enrichment after stratifying by minor allele frequency: variants were separated into 20 genome-wide MAF bins (MAF for a pair was calculated as the average MAF of both groups), a 2×2 table (tested variant set vs all other variants, outlier vs non-outlier) was formed within each bin, and then these tables were combined using the Mantel-Haenszel estimator, resulting in a single odds ratio and its corresponding 95% confidence interval. Significance was evaluated through a one-sided test for enrichment, and p-values were corrected for multiple testing using the Benjamini-Hochberg method across all tests (three genetic ancestry PCs x six group pairs x two variant sets). Analyses were conducted in R (mantelhaen.test).

### Colocalization analysis

To assess colocalization between our eQTLs and other complex traits, we focused on 28 different GWAS across three main categories: autoimmune or inflammatory, hematological, and infectious disease. The traits are listed in the table below.

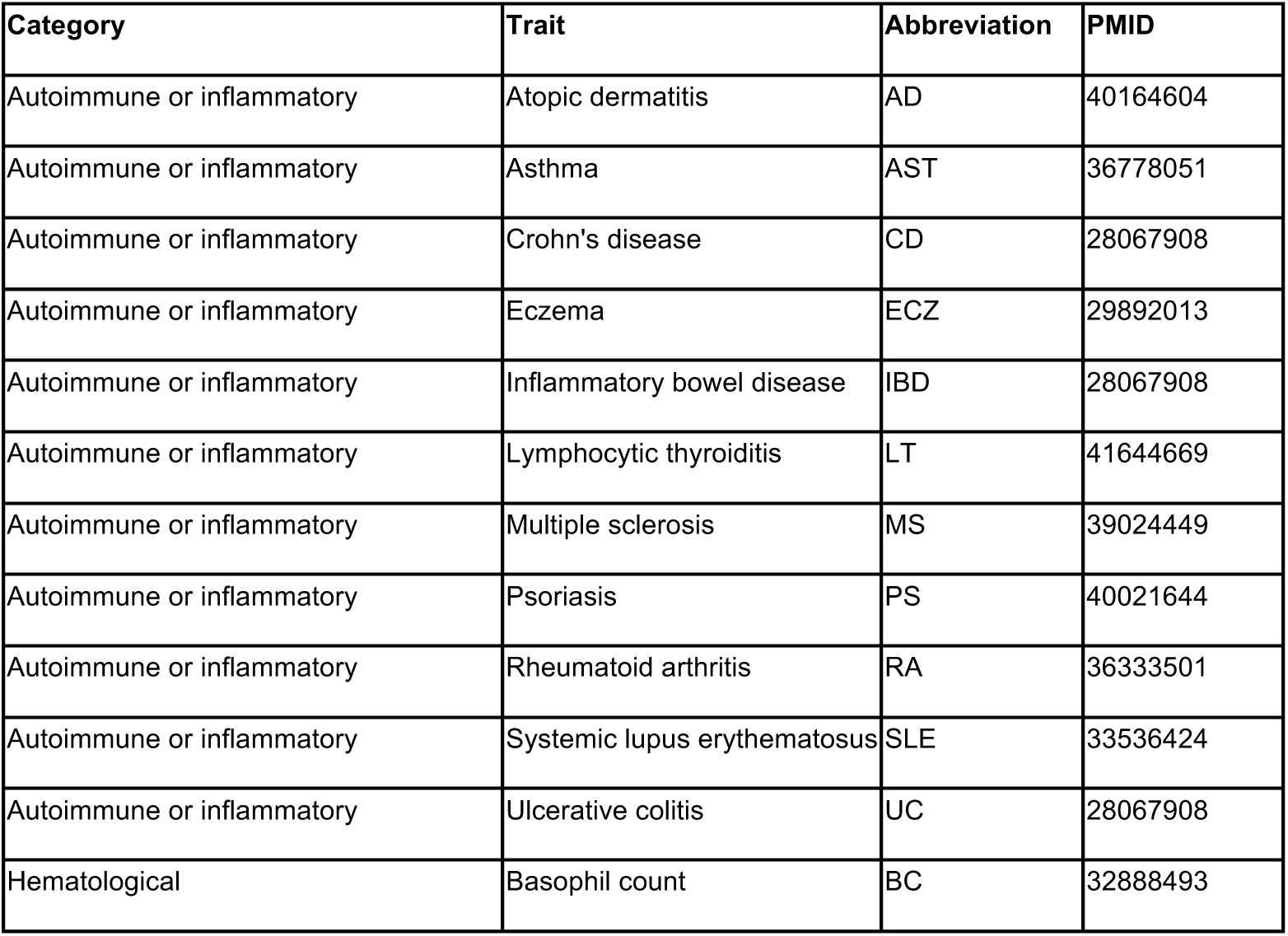
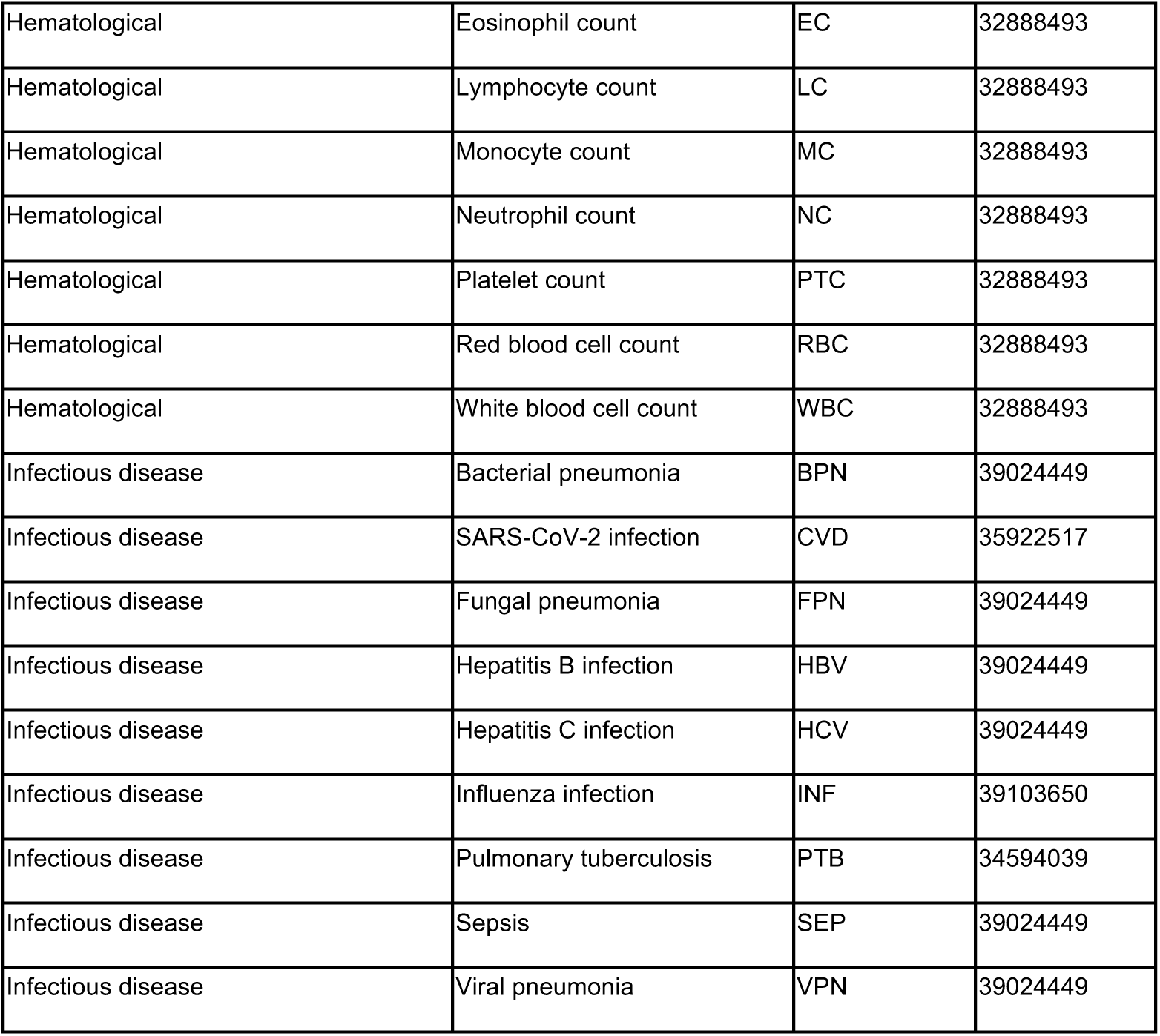

| Category | Trait | Abbreviation | PMID |
| --- | --- | --- | --- |
| Autoimmune or inflammatory | Atopic dermatitis | AD | 40164604 |
| Autoimmune or inflammatory | Asthma | AST | 36778051 |
| Autoimmune or inflammatory | Crohn's disease | CD | 28067908 |
| Autoimmune or inflammatory | Eczema | ECZ | 29892013 |
| Autoimmune or inflammatory | Inflammatory bowel disease | IBD | 28067908 |
| Autoimmune or inflammatory | Lymphocytic thyroiditis | LT | 41644669 |
| Autoimmune or inflammatory | Multiple sclerosis | MS | 39024449 |
| Autoimmune or inflammatory | Psoriasis | PS | 40021644 |
| Autoimmune or inflammatory | Rheumatoid arthritis | RA | 36333501 |
| Autoimmune or inflammatory | Systemic lupus erythematosus | SLE | 33536424 |
| Autoimmune or inflammatory | Ulcerative colitis | UC | 28067908 |
| Hematological | Basophil count | BC | 32888493 |
| Hematological | Eosinophil count | EC | 32888493 |
| Hematological | Lymphocyte count | LC | 32888493 |
| Hematological | Monocyte count | MC | 32888493 |
| Hematological | Neutrophil count | NC | 32888493 |
| Hematological | Platelet count | PTC | 32888493 |
| Hematological | Red blood cell count | RBC | 32888493 |
| Hematological | White blood cell count | WBC | 32888493 |
| Infectious disease | Bacterial pneumonia | BPN | 39024449 |
| Infectious disease | SARS-CoV-2 infection | CVD | 35922517 |
| Infectious disease | Fungal pneumonia | FPN | 39024449 |
| Infectious disease | Hepatitis B infection | HBV | 39024449 |
| Infectious disease | Hepatitis C infection | HCV | 39024449 |
| Infectious disease | Influenza infection | INF | 39103650 |
| Infectious disease | Pulmonary tuberculosis | PTB | 34594039 |
| Infectious disease | Sepsis | SEP | 39024449 |
| Infectious disease | Viral pneumonia | VPN | 39024449 |

For this analysis, we used the coloc R package.^66,67^ For each cell type and condition, we only investigated colocalization for the genes that had a significant eQTL effect, as described in the “Quantitative trait loci mapping” section. To account for multiple colocalization signals, we set the “maxhits” argument of “coloc.signals()” as the number of credible sets identified by susieR or 1 if susieR did not identify any credible sets during fine-mapping. We defined colocalization as PP4 ≥ 0.8.

## Declaration of generative AI and AI-assisted technologies in the manuscript preparation process

During the preparation of this work, the authors used Claude (Anthropic) to assist with generating code to produce figures. The authors also used Claude to refine the language and clarity of the text that the authors had originally drafted. After using this tool, the authors reviewed and edited the content as needed and take full responsibility for the content of the published article.

## Data Availability

All data produced in the present study are available upon reasonable request to the authors.

## Acknowledgements

We thank all study participants and their communities for their invaluable contributions to this work. In Chile, we are grateful to Marcelo Fonseca for valuable contacts; Sue Ellen Rojas for logistical support in participant recruitment; Carolina Ibáñez for facilitating ethics approval at Pontificia Universidad Católica de Chile. From Peru, we thank the following people for their contributions to community engagement and participants recruitment: Marco Fernandez-Manayalle and Carlos Wester (Ministerio de Cultura, Lambayeque, Peru); Amílcar Tinoco (EsSalud Junin Huancayo, Peru); Marisela Montero (Sincos, Peru); Joseph Sulca (USMP, Lima, Peru); Miguel Fernández-Flores (DIGESA-MINSA, Lima, Peru); Carlos Manrique (DIRESA Madre de Dios -MINSA, Puerto Maldonado Peru); Roldán Cárdenas (DIRESA Loreto-MINSA, Iquitos, Peru); Víctor Giú (DIRESA Ucayali -MINSA, Pucallpa, Peru). We thank members of the Barreiro lab for their constructive comments and feedback. We thank Haley Randolph for helpful clarifications regarding her publicly available code, and Tauras P. Vilgalys for critical reading of the manuscript. This work was completed in part with resources provided by the University of Chicago Research Computing Center, and we thank the University of Chicago Genomics Facility (RRID: SCR_019196) for their assistance with sequencing. This work was supported by the Chan Zuckerberg Initiative Foundation (DAF2022-240351) [to L.B.B., L.V., R.F., and T.H], R35-GM152227, and P01-AI189645 [to L.B.B.], and by ANID FONDECYT grant 11200324 [to L.V.]. The funding agencies had no role in the design, implementation, or interpretation of this study.

## Author Contributions

L.B.B. and L.V. conceived and designed the study. L.B.B. supervised the study. L.V., T.H., R.F., and L.B.B. acquired funding. Community engagement and recruitment were performed by L.V., Y.F., P.B., and J.Bo. in Chile, by R.F. and O.A. in Peru, and by J.Gu. and T.H. in Brazil. J.Go., J.L., and T.C. collected blood samples from the Chilean populations. D.O., M.L.G.-F., S.F., C.A., J.R.G., T.L., T.R., A.K., C.C., M.P., B.C., A.D., K.W., and L.B.B. isolated and cryopreserved PBMCs from the Peruvian, Chilean, and Brazilian samples. DNA extractions were performed by L.V., C.C., M.L.G.-F., D.O., M.P., J.R.G., and T.L. Single-cell experiments and infections were primarily performed by C.C., M.P., C.J., and K.Me., with contributions from J.Ba., L.V. and F.C. K.Ma. and I.F. provided instrumentation and laboratory infrastructure. R.L. and C.K.M. provided influenza virus preparations, and E.S. and D.S. provided *T. cruzi* preparations. C.C., L.V., and L.B.B. analyzed the single-cell and genetic data. D.A. performed the eQTL and colocalization analyses, E.W. performed the F_ST_ analyses, A.G. performed genotype imputation, and R.A.-G. contributed to computational analyses and figures. B.C. contributed pipelines for the analysis of genetic data. L.V., C.C., and L.B.B. primarily wrote the manuscript, with contributions from D.A., and E.W. All authors provided feedback on the manuscript.

---

**Figure S1.**
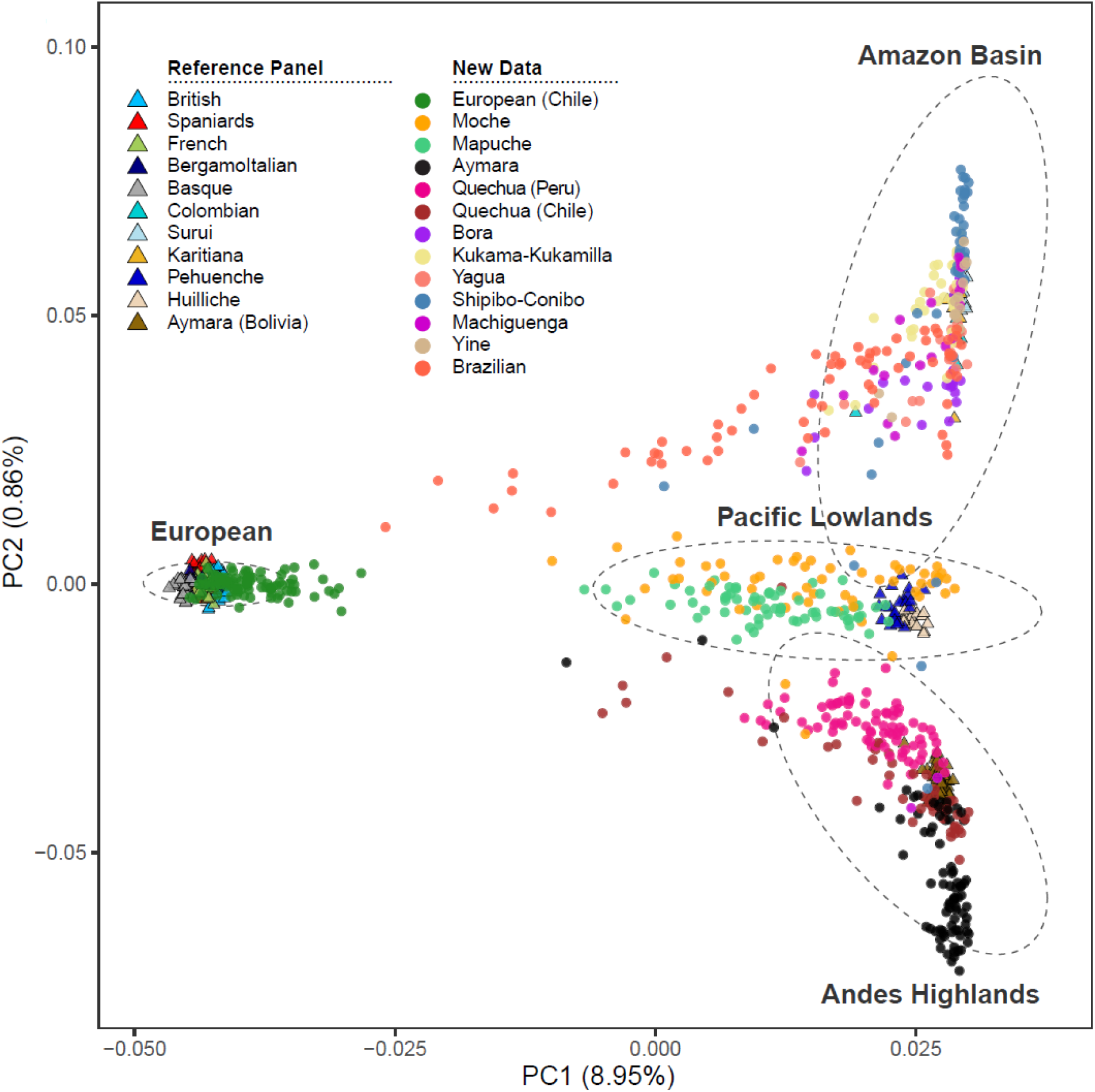
Principal component analysis on genotype data of reference populations and the new data from this study.

**Figure S2.**
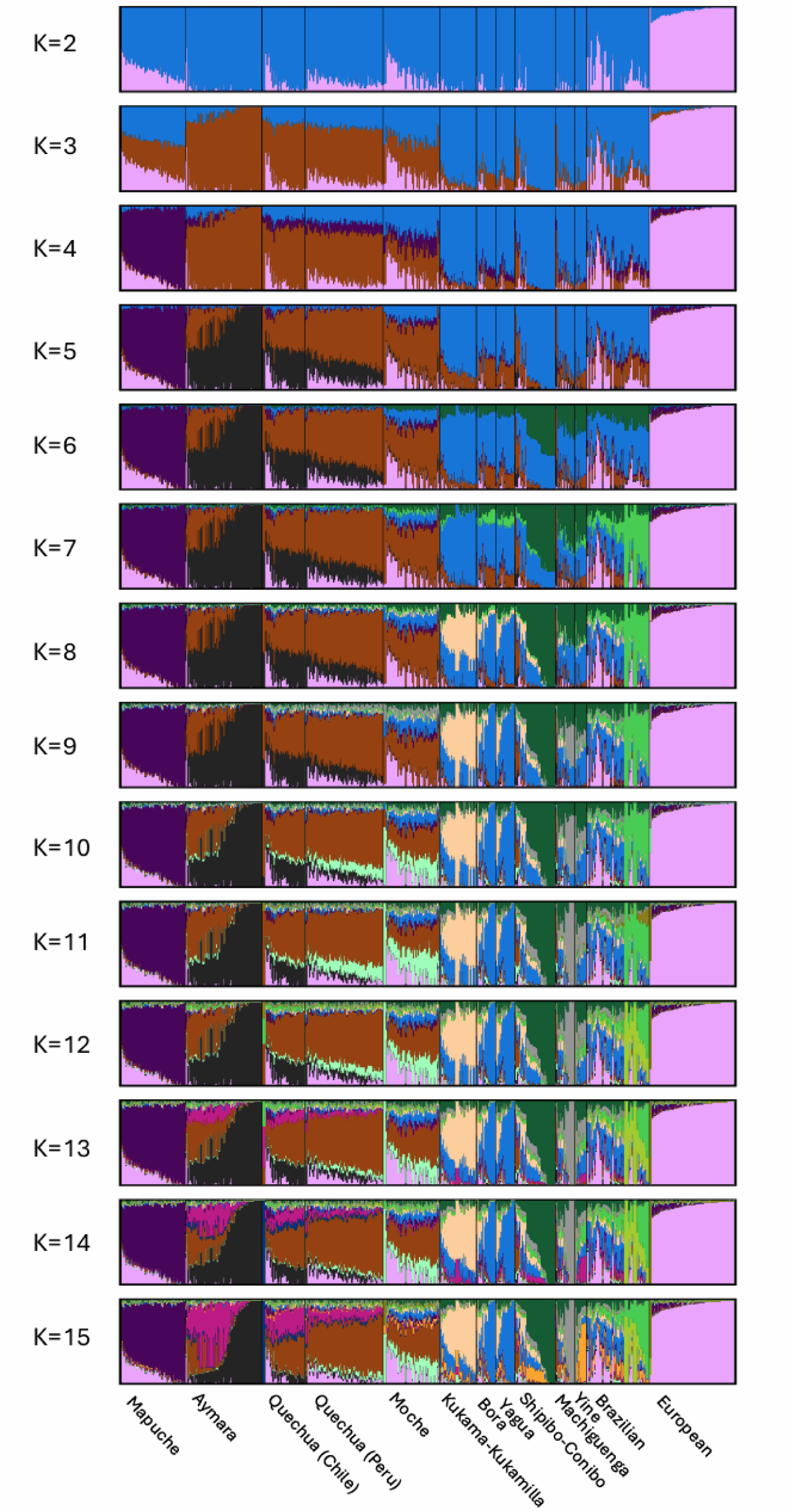
ADMIXTURE results at *K = 2* through *K = 15*.

**Figure S3.**
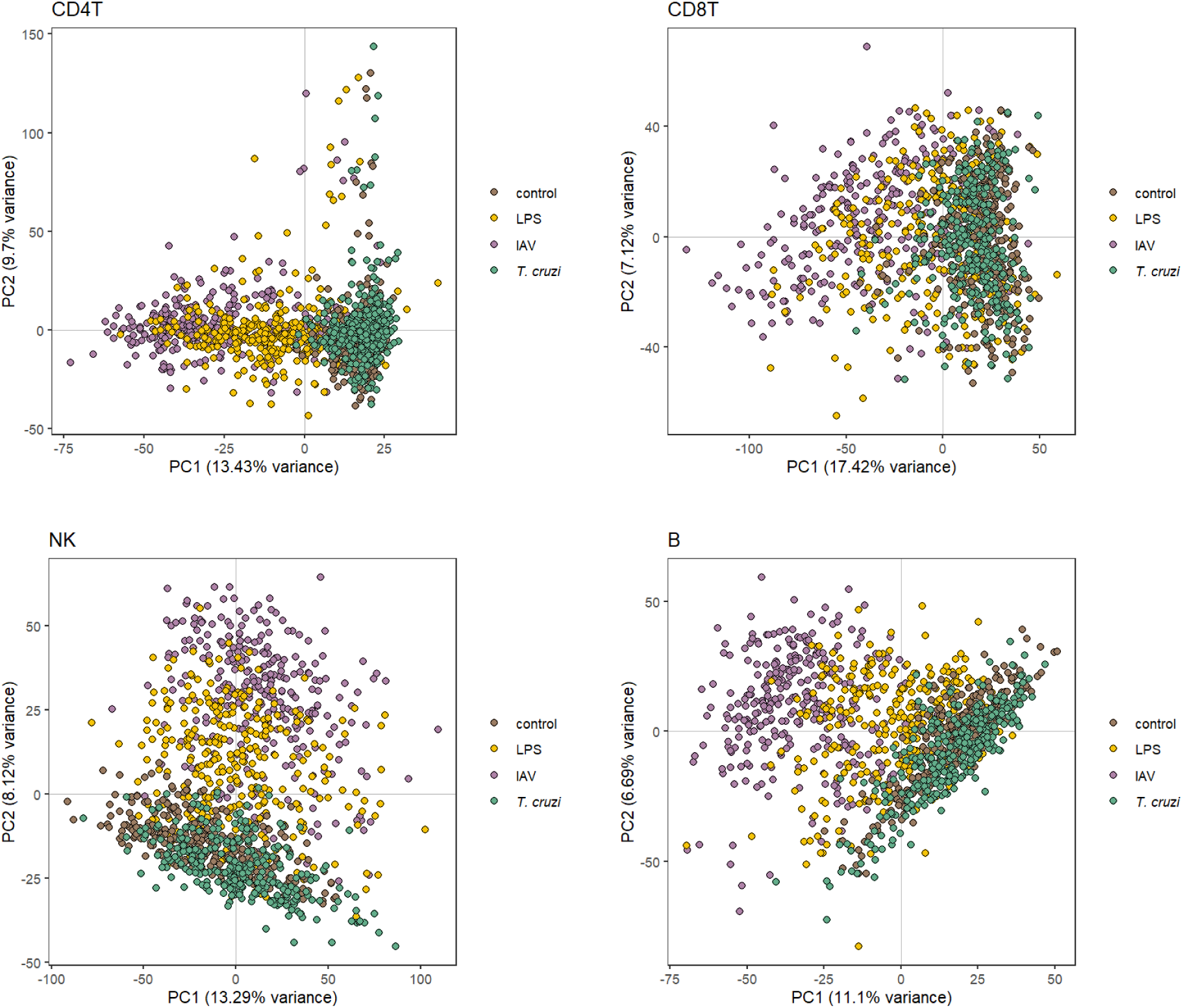
Principal component analysis on pseudobulk gene expression matrices of other PBMC cell types.

**Figure S4.**
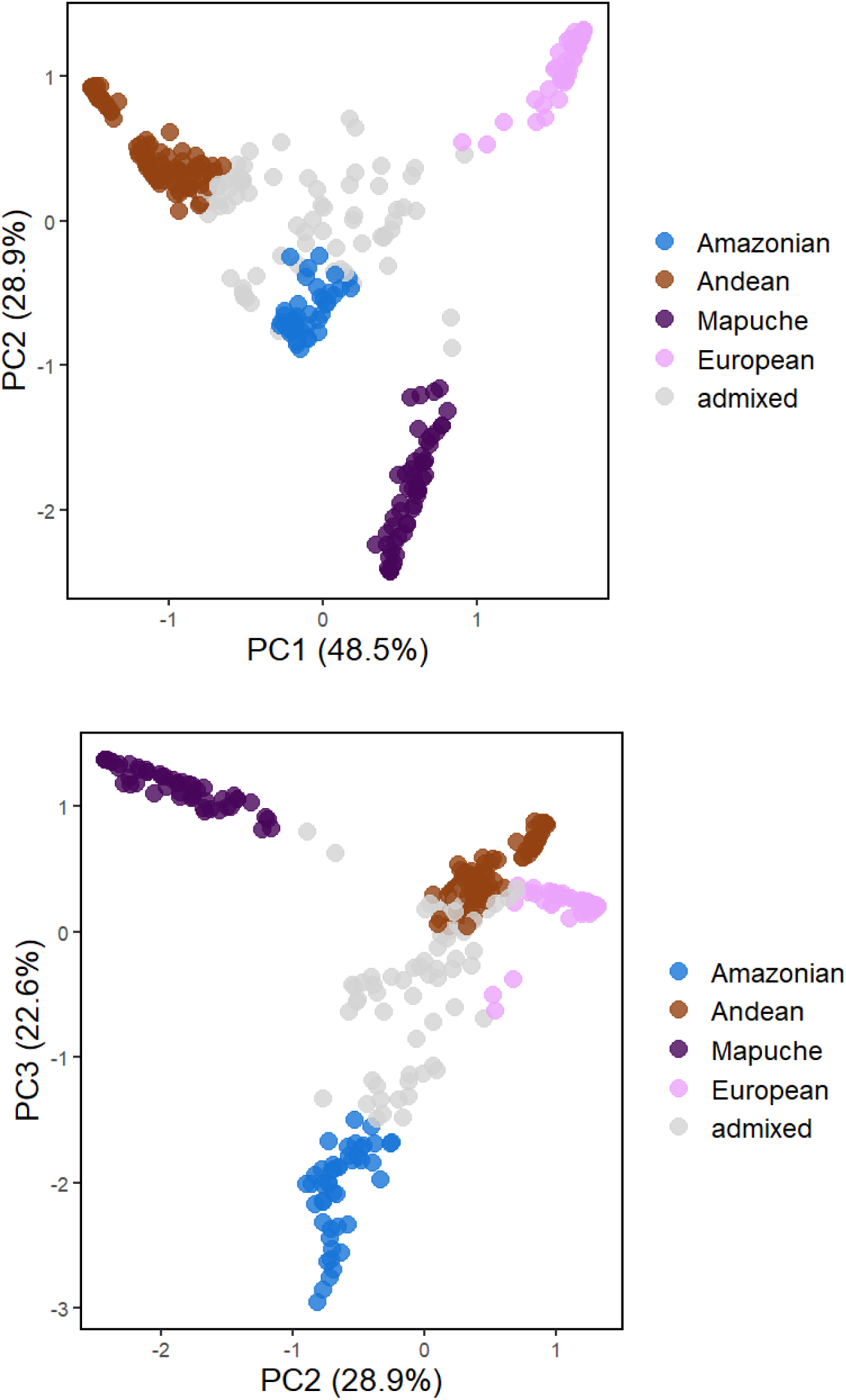
Principal component analysis of ADMIXTURE ancestry proportions at *K = 4*.

**Figure S5.**
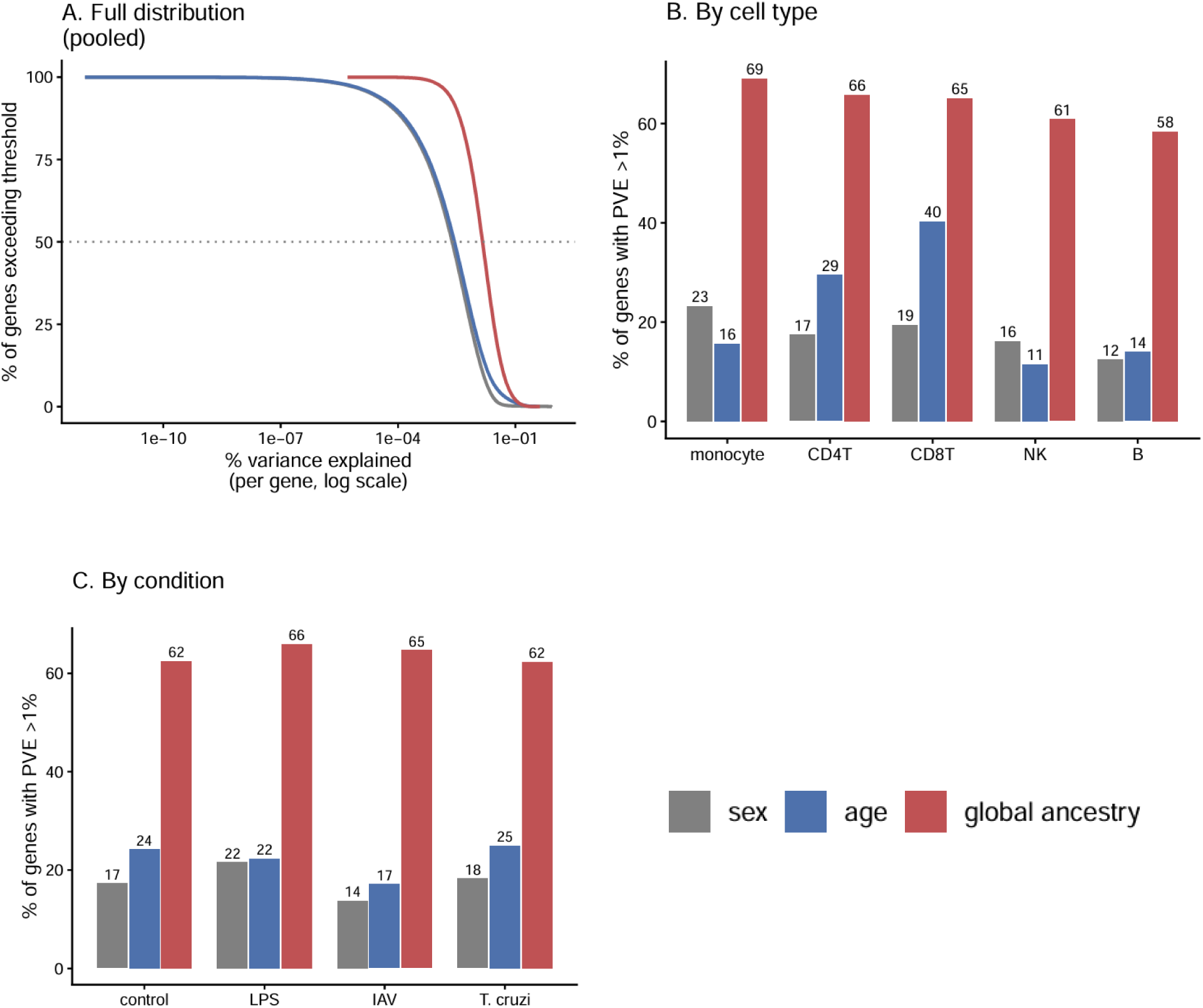
Percentage of genes with percent variance explained (PVE) > 1% by cell type and condition grouped by sex, age, and global genetic ancestry.

**Figure S6.**
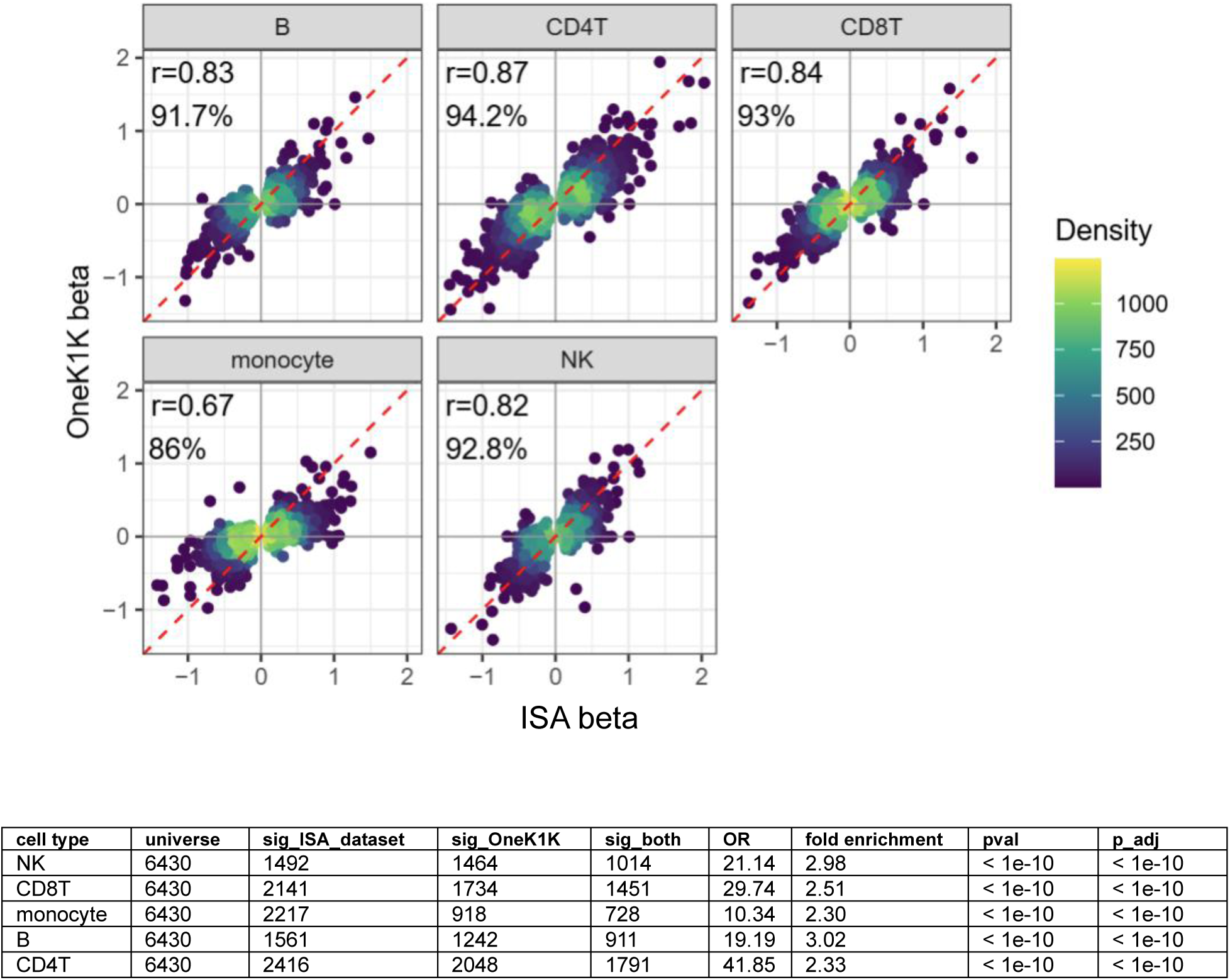
Correlation of OneK1K eQTL effect sizes (betas) with our ISA dataset. Table shows results of Fisher’s exact test.

**Figure S7.**
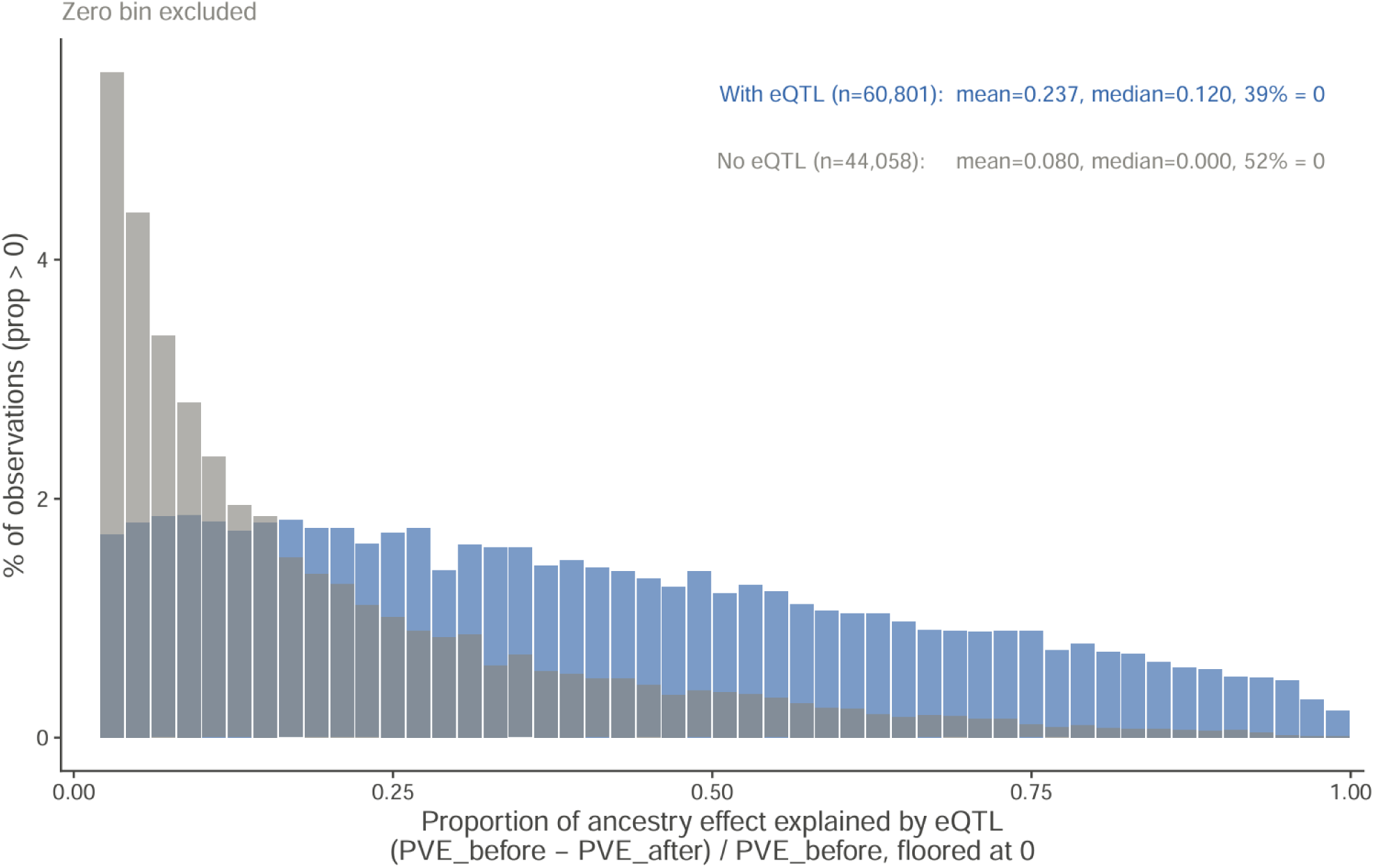
Distribution of observations (unique gene-condition-cell type combinations) by proportion of ancestry effect explained by independent eQTL signals.

**Figure S8.**
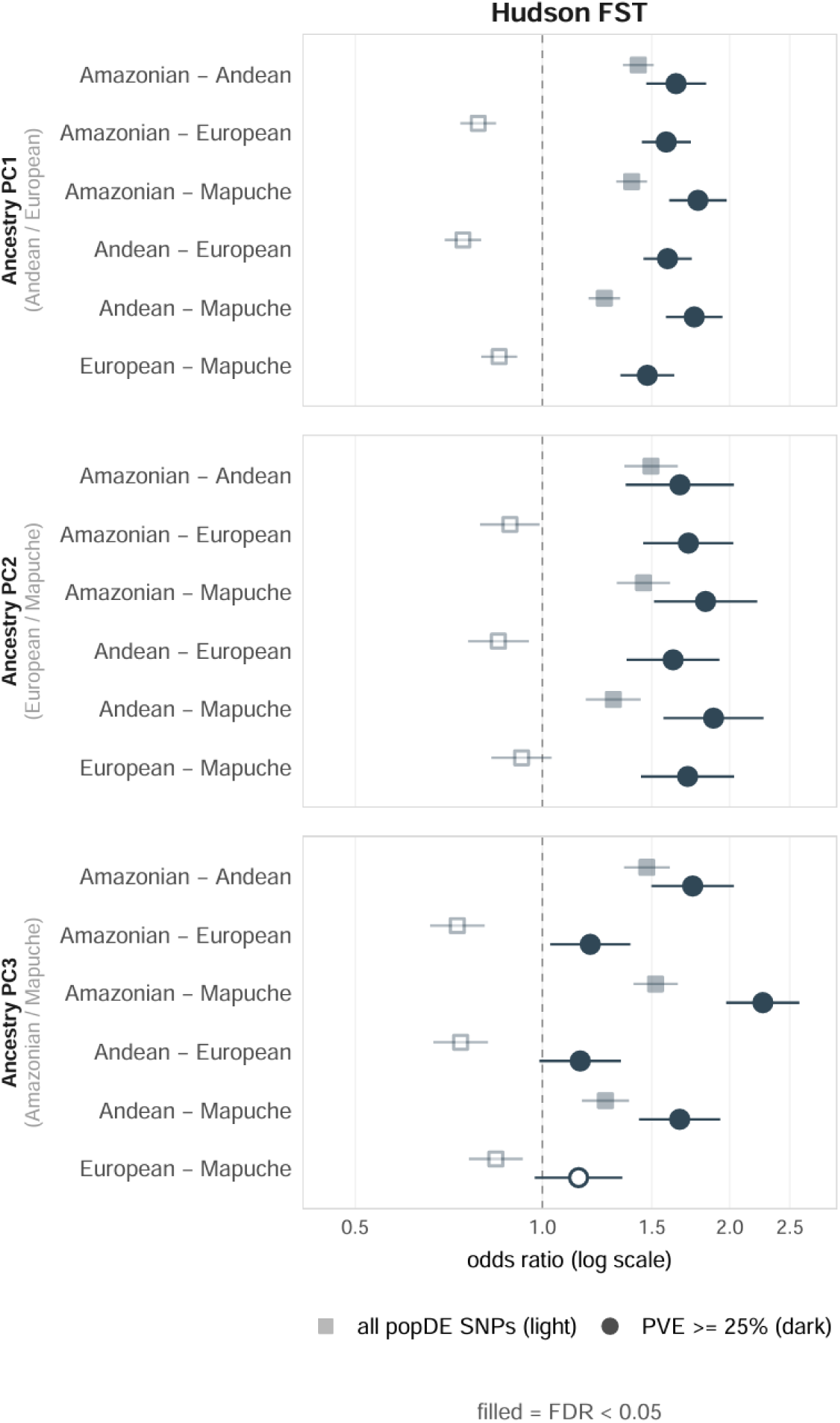
Odds ratios for enrichment of eQTLs associated with popDE genes among genome-wide F_ST_ outliers.

**Figure S9.**
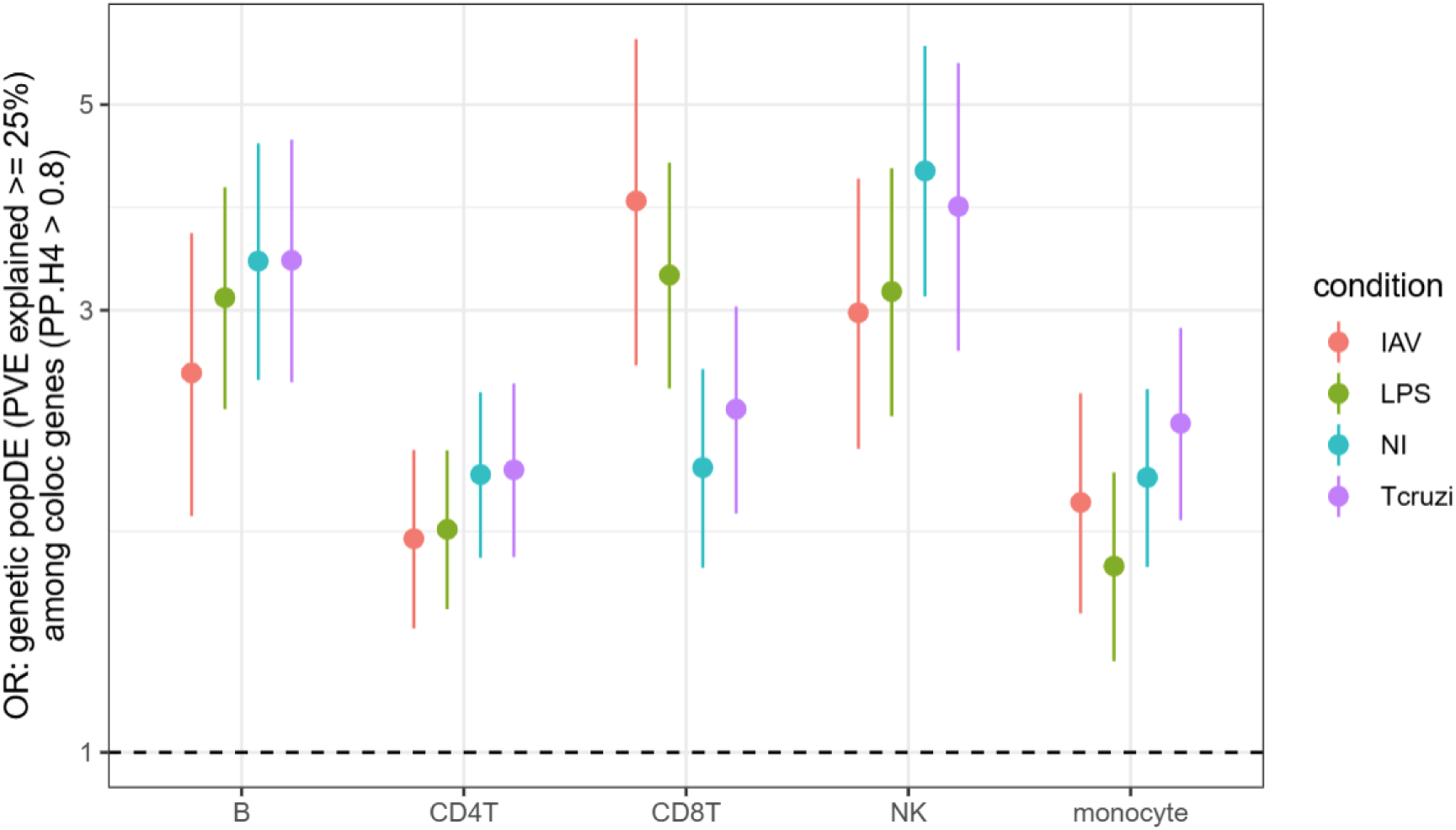
Enrichment of popDE genes (with eQTLs explain at least 25% of the ancestry-associated variance in gene expression) among genes colocalizing with at least one of 28 immune-related GWAS traits (PP.H4 > 0.8).

